# *TGFB1* rs8179181 polymorphism is reproducibly associated with Parkinson’s disease in a Spanish population

**DOI:** 10.1101/2022.07.09.22277447

**Authors:** Alicia Comino, Mónica Antolín-Vallespín, Azahara López-Benito, Gloria Muñoz, Francisco Javier del Castillo, Lydia Vela, Juan Carlos Martínez-Castrillo, Amelia Sánchez-Capelo

## Abstract

There is evidence that transforming growth factor (TGF)-β signaling participates in the pathology of Parkinson’s disease (PD). Dampened TGF-β signaling in Smad3- or TβRII-deficient mice leads to the appearance of α-synuclein inclusions in the brain, as well as dopaminergic, motor, and cognitive deficits. Accordingly, we hypothesized that genetic variants of *TGFB*/*SMAD* could be risk factors for PD in humans. Here, we present two independent case-control studies aimed at evaluating the association between genetic variants of six genes related to TGF-β signaling (*TGFB1, TGFB2, TGFBRI, TGFBRII, SMAD3* and *SMAD2*) and the development of sporadic PD. A total of 275 unrelated Spanish Caucasian individuals were included in the study (141 cases and 134 controls), with 132 individuals in the discovery phase and 143 individuals in the replication phase. Next-generation sequencing identified a total of 409 variants in the coding, splicing, and untranslated regions of these genes. Analysis of common variants in the discovery phase revealed an association between PD and the *TGFB1* rs8179181 variant, which was further confirmed in the replication phase [odds ratio (OR) 0.48, 95% confidence interval (CI) 0.32–0.73, p = 0.00057). A weak association of the *SMAD3* rs11556089 polymorphism with PD was also detected (OR 0.49, 95% CI 0.26–0.93, p = 0.0375). Seven haplotypes were identified; however, there were no significant differences in their frequencies between patients with PD and controls. In conclusion, both the discovery and replication phases of this study suggest that the rs8179181 variant of *TGFB1* represents a novel susceptibility locus for PD.

**HIGHLIGHTS:**

- Deficient TGF-β signalling via Smad3 induces the formation of α-synuclein aggregates and a parkinsonian pathology in mice.
- *TGFB1, TGFB2, TGFBRI, TGFBRII, SMAD3* and *SMAD2* genes were sequenced to search for associations of their allelic variants with idiopathic PD.
- Two independent case-control studies of Spanish Caucasian individuals identified 409 genetic variants in their coding, splicing and UTR regions.
- The rs8179181 SNP in *TGFB1* was reproducibly associated with idiopathic PD, representing a novel PD susceptibility locus.

## INTRODUCTION

With the aging of the global population, Parkinson’s disease (PD) is becoming an increasingly important disease and its prevalence is expected to double over the next two decades [1]. PD is diagnosed as a progressive movement disorder, although non-motor symptoms are also frequent and potentially disabling [2]. Neuropathically, PD is characterized by slow neurodegeneration of the dopaminergic neurons in the substantia nigra (SN) and accumulation of α-synuclein aggregates [3]. Among the factors considered to be involved in the development of PD, transforming growth factor-beta (TGF-β) signaling has been implicated in distinct pathological events related to PD, including the development of α-synuclein inclusions [4, 5].

TGF-β is a large superfamily of extracellular molecules encoded by 33 genes in mammals, which is mainly subdivided into the TGF-β, bone morphogenic protein (BMP), and growth differentiation factor (GDF) families. The TGF-β family signals through a canonical intracellular transduction pathway that is triggered by the interaction of extracellular TGF-β1, TGF-β2, and TGF-β3 ligands with the TβRI and TβRII membrane receptors, thereby promoting the phosphorylation and nuclear translocation of Smad2/3 transcription factors [6]. TGF-β1 is a pleiotropic molecule that regulates cell proliferation, migration, and differentiation. Indeed, its effects are cell type- and cell context-dependent, and may provide signals for both cell survival and apoptosis [7].

TGF-β1 is chronically upregulated in striatal regions and in the ventricular cerebrospinal fluid of patients with PD [8, 9], similar to other brain disorders such as Alzheimer’s disease (AD) [10, 11]. In adult mice, TGF-β1/-β2, TβRI, TβRII, Smad2, and Smad3 are expressed in both the SN and the striatum. Moreover, studies with experimental animal models have suggested that chronic TGF-β1 overexpression may contribute to the pathology of PD [12-15], and deficient TGF-β signaling may be a risk factor for developing parkinsonism [16-19]. Smad3 deficiency promotes α-synuclein and ubiquitin inclusions that adopt a core/halo distribution resembling human Lewy bodies (LBs). This deficiency also promotes postnatal dopaminergic neurodegeneration, strong MAO-mediated catabolism of dopamine, and dampened trophic and astrocyte support for dopaminergic neurons [17]. Similarly, inhibition of the TβRII receptor promotes dopaminergic neurodegeneration and motor deficits [18]. Smad3 deficiency can also induce cognitive deficits as it enhances GABA_A_ neurotransmission, leading to the inhibition of long-term potentiation (LTP) and diminishes neurogenesis in the adult hippocampus [20, 21]. Indeed, TβRII-deficient mice have enhanced GABAergic input to the dopaminergic neurons, provoking hyperactivity and reversal learning deficits [22].

Genetic factors that follow Mendelian inheritance suggest that familial PD arises in only 10% of PD patients. In contrast, idiopathic PD is a complex multifactorial disorder caused by interactions between genetic and environmental risk factors, where common genetic variants have small effects [23]. Genome-wide association studies (GWAS) have identified several common variants that may be considered risk factors for PD; however, most of these GWAS-identified variants have not been assessed in functional assays, and the majority of the heritable component of the disease remains to be fully defined [23-25]. Next-generation sequencing (NGS) can fine-map target genes, as opposed to the tagging of selected single nucleotide polymorphisms (SNPs) in GWAS approaches. Interestingly, NGS can complement GWAS in the discovery of additional risk loci through association studies of candidate genes, thereby identifying novel variants that are not otherwise detected in GWAS [26].

Considering the implications of TGF-β signaling in experimental models of PD, individuals with altered TGF-β signaling may be at risk of developing this disease. Indeed, a genetic association study showed that the haplotype-tagged SNP rs6658835, located in the 5′ region of the *TGFB2* gene, is associated with susceptibility to PD [27]. In this study, we used NGS to search for allelic variants in the exons, untranslated regions (UTRs), and intronic splice regions of six genes in the TGF-β signaling pathway (*TGFB1, TGFB2, TGFBR1, TGFBR2, SMAD2* and *SMAD3*). Two independent case-control studies were conducted on a cohort of unrelated Spanish Caucasian individuals. As a result, we identified a new association of the rs8179181 variant of *TGFB1* with idiopathic PD.

## PATIENTS AND METHODS

### Study design and cohort

Two independent case-control studies were carried out on 275 unrelated Spanish Caucasian individuals to study the association of PD with genetic variants in six TGF-β genes: *TGFB1, TGFB2, TGFBRI, TGFBRII, SMAD2*, and *SMAD3*. The first association study (discovery phase) included 70 patients with idiopathic PD (44 men and 26 women) and 62 neurologically healthy controls (37 men and 25 women) (**Table 1**). The second study was an exact replicate of the first study (replication phase) including 71 patients with idiopathic PD (38 men and 33 women) and 72 neurologically healthy controls (27 men and 45 women). Thus, the final dataset (joint analysis) included 141 patients with idiopathic PD (82 men and 59 women) and 134 neurologically healthy controls (64 men and 70 women). This sample size offered 80% power for detecting a disease variant with a risk ratio of 1.80 in a two-stage design [28]. The time frame between the discovery and replication phases was 18 months, and the participants were recruited using different strategies, with 253 individuals recruited *via* the Carlos III Spanish National DNA Bank (BNADN: www.bancoadn.org, Salamanca, Spain) from donors attending hospitals in 13 different regions across Spain. In addition, 22 participants were recruited from the Hospital Universitario Ramón y Cajal and Hospital Universitario de Alcorcón (both located in Madrid). The mean age of the PD patients at disease onset was 63.1 ± 0.6 years (range 48–81 years) and the mean age of the controls at recruitment was 62.5 ± 0.2 years (range 48–78 years; P = 0.690). PD diagnosis was made based on established criteria [29, 30], and all patients were included in the study irrespective of disease severity. Participants who reported a family history of PD (from both the patient and control groups) were excluded from the study. The age at onset was defined as the age at which the patient noticed the first symptom indicative of PD. Neurologically healthy controls were recruited among individuals living in the same geographical area as the patients who visited the clinics and were free of any neurological disease or stroke. As nicotine has a potential neuroprotective effect, we also excluded smokers from the study or participants who smoked in the previous 20 years [31]. After obtaining approval from the hospitals’ Ethical and Scientific Committees and informed consent from the subjects, blood samples were drawn for DNA extraction.

**Table 1.**
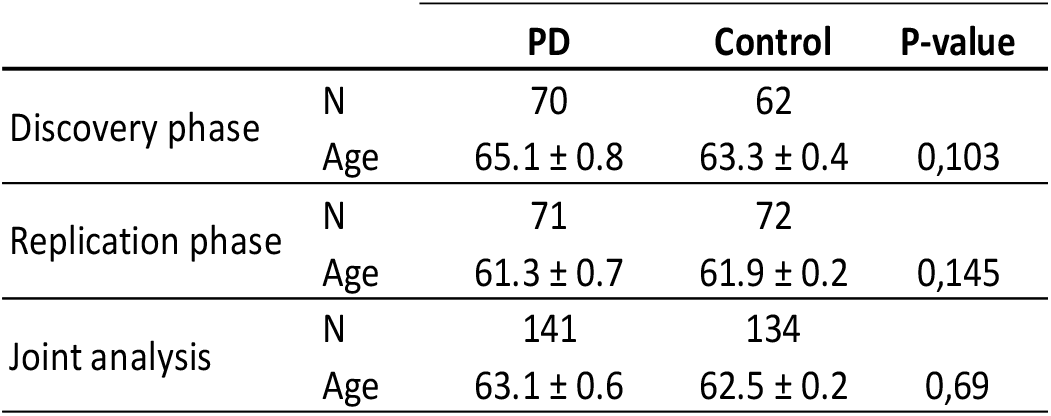
Characteristics of the study population. The data are shown as the mean ± SEM: N, number of individuals; Age, age at onset of PD in patients and age at blood drawing in controls.

### Ion Torrent sequencing

Genomic DNA was extracted from peripheral blood collected in EDTA using the QIAamp DNA Blood kit (Qiagen, Germany), determining the DNA concentration with the Qubit™ dsDNA BR Assay kit (Invitrogen, Life Technologies, USA) and Qubit 2.0 Fluorometer (Invitrogen). Genomic DNA was analysed by Ion Torrent NGS at the Central Translational Genomics Research Facility of the Hospital Ramón y Cajal (UCA-GT). We performed NGS of exons, exon/intron boundaries and UTR regions of six genes in the TGF-β intracellular signalling pathway: *TGFB1* (GenBank NM_000660), *TGFB2* (NM_001135599), *TGFBR1* (NM_001130916), *TGFBR2* (NM_003242), *SMAD2* (NM_005901), and *SMAD3* (NM_005902). The selection of these genes allows allelic variants in the canonical intracellular signalling pathway of the TGF-β family to be fine mapped [6]. NGS was performed using an AmpliSeq strategy on an Ion Torrent personal genome machine (PGM) platform (Life Technologies) and the primer sets used were obtained using Ion AmpliSeq™ Designer (v3.0.1, Applied Biosystems, Life Technologies). A total of 236 amplicons were designed to cover 42.23 Kb, an overall coverage of 91.25% of the coding sequences, splice, and 5’-, 3’-untranslated regions of the six TGF-β genes, plus the promoter of Smad3 (for the exact coordinates and primer sequences see **Supplementary Material S1** and **S2**).

#### Amplicon Library Preparation

Initial amplification of the targeted regions was performed using the Ion AmpliSeq™ Library Kit 2.0 and Ion Xpress™ Barcode Adapter 1–16 Kit (Applied Biosystems), according to the manufacturers’ instructions but using half the volumes. PCR amplification was performed on 10 ng of template DNA, with 2 µl of the 5X Ion AmpliSeq™ HiFi Master Mix (Applied Biosystems) and 2 µl of the 5X Ion AmpliSeq™ Custom primer pools, employing the following PCR protocol (GenAmp PCR system 9700, Applied Biosystem): 1 cycle at 99 °C for 2 min; 18 two-step cycles at 99 °C for 15 s and 60 °C for 4 min; a final holding period at 10 °C. After multiplex PCR amplification, the DNA amplified was digested for 10 min at 50 °C and for 10 min at 55 °C with the FuPa enzyme (1 µl), which was then inactivated at 60 °C for 20 min. After digestion, 4 µl of the diluted barcode adapter mix (which included the Ion Xpress™ Barcode Adapter and Ion P1 adapter) was ligated to the end of the digested amplicons for 30 min at 22 °C, with 2 µl of the Switch solution and 1 µl of DNA ligase, after which the ligase was inactivated at 72 °C for 10 min. The adaptor ligated amplicon libraries were cleaned up using the Agencourt AMPure XP beads (Beckman Coulter Genomics, USA), from which they were eluted and then amplified using 25 µl of the Platinum PCR SuperMix High Fidelity with 1 µl of the Library Amplification Primer Mix. The amplified libraries were purified following a two-round purification process with the Agencourt AMPure XP system, and the purified libraries were quantified with the Qubit™ dsDNA HS Assay kit (Invitrogen) using a Qubit 2.0 Fluorometer (Invitrogen). A Tape Station instrument with the Agilent High Sensitivity D1000 Screen Tape was used to analyse the profile of the libraries and after quantification, each amplicon library was diluted to 100 pM. Sequencing Chip316v2 was used and for each chip, the template DNA was pooled such that each pool contained equimolar amounts of DNA from 16 individuals.

#### Emulsion PCR and Sequencing

The pools of libraries were then diluted to a concentration of 30 pM and emulsion PCR was performed using the OneTouch™ 2 System and the Ion OneTouch 200 Template Kit v2 (Life Technologies), according to the manufacturer’s instructions. Template-positive Ion Sphere™ particles were enriched with Dynabeads MyOne™ Streptavidin C1 and washed with Ion OneTouch™ Wash Solution using the Ion OneTouch™ ES system (Life Technologies). After Ion Sphere Particle preparation, the sequencing 316™ chips were loaded and sequencing was performed on an Ion Torrent PGM system using Ion PGM™ 200 Sequencing Kits (Life Technologies) according to the established procedures.

#### Base call and data analysis

The sequence data were processed using the Ion Torrent Suite™ 4.0 software and the Torrent Server. The sequences were aligned against the human reference sequence (build GRCh37) using a Torrent Mapping Alignment Program optimized to Ion Torrent data, trimming away the primer sequences. The reads were aligned against the designed BED files to obtain the FASTQ data and the alignments were visually checked with the IGV software (Integrative Genomics Viewer, https://software.broadinstitute.org/software/igv/). Variant calling was then performed with the Variant Caller plug-in software, using parameters tuned for low stringency. Coverage analysis was performed using the Coverage plug-in from the Ion Torrent software Suite. Annotation of the variants was accomplished with the UCSC and Ensemble genome browsers (https://genome-euro.ucsc.ed, https://www.ensembl.org), dbSNP v153 (https://www.ncbi.nlm.nih.gov/snp) and from publications in the PubMed database (https://www.ncbi.nlm.nih.gov).

### Genotyping and Quality Control Assessment

Quality scores were determined for each single nucleotide variation (SNV) using Ion Torrent Suite 4.0. We manually curated all genetic data through visual inspection using IGV software. As such, we performed two rounds of manual genotype calling, in which we obtained every SNV present in the six TGF-β genes for each patient. In the second round, we verified each SNV by considering the whole study cohort. Each SNV had to fulfill the following criteria for the manual genotype calling of variants: Phred score >20, mapping score > 30, variant present on both forward and reverse strands, and heterozygosity when the variation was present in 20–80% of the reads. We performed further overall quality control analysis using PLINK software v.1.07, excluding markers with the following characteristics: a missing genotyping rate > 5%, a minor allele frequency (MAF) < 1%, and deviation from Hardy-Weinberg equilibrium (HWE) at p < 0.001 in the controls [32].

In association studies of candidate genes, the most efficient way to determine population stratification is to use ancestry-informative markers (AIMs) [33, 34]. We searched for AIMs in the six TGF-β genes using the 1000G Phase 3 database. We obtained allele frequencies for the Iberian population in Spain (IBS) and the European Caucasian population (EUR), which were compared with those of other populations such as the African (AFR), South Asian (SAS), East Asian (EAS), and Ad-Mixed American (AMR) populations. We selected SNVs with the maximum capacity to differentiate EUR populations.

### Statistical Analysis

For common variants with a MAF ≥ 1%, single-variant PD association analysis was performed, estimating the odds ratios (ORs) and 95% confidence intervals (CIs) with PLINK. The association was tested using logistic regression analysis adjusted for the covariates of sex and age, using both the individual datasets of the discovery and replication phases, as well as the pooled data for the joint analysis. In addition, we performed haplotype construction and haplotype association analyses using logistic regression adjusted for the same covariates. The max(T) permutation approach was used in both allele-based and haplotype-based analyses to correct for multiple testing, obtaining a p-value adjusted through 100,000 permutations. Permutation is the method of choice for accurate correction in candidate gene association studies [34-36]. CaTS software was used to calculate the power of the two-stage association study [28].

## RESULTS

### NGS and variant analyses

Ion Torrent sequencing of the coding, splicing, and untranslated regions of six TGF-β related genes was performed on 132 individuals in the discovery phase and 143 individuals in the replication phase, providing data from 275 individuals for the joint analysis (**Table 1**).

Nineteen sequencing chips were used, obtaining average reads of 3.34 Gb per chip, and the sum of the overall chips produced a total read of 60.10 Gb of DNA sequence, with 93.4% of the reads at AQ20. The mean number of total reads for each amplicon and individual was 792 ± 55. Only 10 out of 236 amplicons did not achieve an average of 20× coverage per amplicon, and none of the individuals failed to achieve 75× coverage on average. After read mapping, SNVs were detected with either Ion Torrent software or by manually curating the data collected. A comparison between the SNVs obtained from the genotype calling algorithms and the manual curation indicated that calling algorithms produced 16% false-positive and 12% false-negative genotypes relative to the manually curated data, which are in line with values obtained using similar algorithm systems [37-39]. The false-positive/negative data generated by the algorithms were dependent on the gene and type of SNV, and were more frequent for insertions/deletions than SNPs (data not shown). In summary, the combined algorithm and manual calling approach improved the quality of the dataset, from which 409 genetic variants were identified (**Supplementary Material S3**). Of these, 115 were new variants (not present in dbSNP v153), 46 were present in coding regions, 29 were in 5’ UTRs, 60 in introns, 254 in 3’ UTRs, and 20 were upstream or downstream of the genes (**Table 2**).

**Table 2.** Number of variants detected in the 275 Spanish individuals: ^1^ Number of base pairs read per individual; ^2^ Variants not present in dbSNP v153.

| Gene | Base-pairs<sup>1</sup> | Total | % | New<sup>2</sup> | Coding region | Upstream | 5'UTR | Intron | 3'UTR | Downstream |
| --- | --- | --- | --- | --- | --- | --- | --- | --- | --- | --- |
| *TGFB1* | 3463 | 32 | 0.92% | 7 | 11 | 0 | 4 | 5 | 0 | 12 |
| *TGFB2* | 5758 | 73 | 1.27% | 24 | 8 | 0 | 16 | 10 | 39 | 0 |
| *SMAD3* | 7514 | 99 | 1.32% | 28 | 11 | 6 | 7 | 25 | 49 | 1 |
| *SMAD2* | 10278 | 90 | 0.88% | 30 | 2 | 0 | 0 | 11 | 77 | 0 |
| *TGFBR1* | 6373 | 72 | 1.13% | 17 | 5 | 0 | 0 | 6 | 61 | 0 |
| *TGFBR2* | 4581 | 43 | 0.94% | 9 | 9 | 1 | 2 | 3 | 28 | 0 |
| Total | 37967 | 409 | 1.08% | 115 | 46 | 7 | 29 | 60 | 254 | 13 |

To evaluate population stratification, we searched for a set of variants for AIMs with the maximum ability to differentiate the EUR population. We found three SNPs (rs8179181 in *TGFB1*, and rs11638476 and rs8028147 in *SMAD3*) with different allele frequencies in the IBS and EUR populations than in the other populations. The recruitment of patients focused on a Spanish Caucasian population, and the MAF values for these three AIMs in our local dataset were similar to those in the IBS and EUR populations in the 1000 Genomes database (**Table 3**), confirming that the study population was of European Caucasian ethnicity.

**Table 3.** AIMs identifying the European population. MAFs obtained from 1000G Phase 3 database: Spanish (IBS), European Caucasian (EUR), African (AFR), South Asian (SAS), East Asian (EAS), and Ad Mixed American (AMR). ^1^Chromosome position (chromosome:bp position) according to the NCBI database, build GRCh37.

| Gene | SNV ID | Position<sup>1</sup> | Alleles | local | IBS | EUR | AFR | SAS | EAS | AMR |
| --- | --- | --- | --- | --- | --- | --- | --- | --- | --- | --- |
| *TGFB1* | rs8179181 | 19:41838206 | C/T | 0.253 | 0.271 | 0.243 | 0.005 | 0.059 | 0.004 | 0.099 |
| *SMAD3* | rs11638476 | 15:67485101 | G/A | 0.367 | 0.453 | 0.426 | 0.089 | 0.147 | 0.001 | 0.233 |
| *SMAD3* | rs8028147 | 15:67418233 | G/A | 0.271 | 0.215 | 0.211 | 0.428 | 0.470 | 0.613 | 0.416 |

Quality-control filtering indicated that none of the individuals had low genotype rates (<95%). Briefly, in the pooled data (joint analyses), we excluded 14 variants with a high missing rate (>5%), whereas the call rate did not differ between PD cases and controls for any of the variants identified. Of the variants analyzed, 216 had a MAF <1%; accordingly, they were considered rare variants and were not studied further. Finally, 16 variants that failed the HWE test in controls were excluded, leaving 163 variants to be studied in more detail.

### Single variant disease-association analysis

An association analysis was performed using logistic regression, adjusting for the covariates of age and sex to avoid any survival and gender bias. The genomic inflation factor (λ) was 0.959, indicating no substantial inflation of the test statistics. In the initial association study (discovery phase), the strongest SNP signal was obtained for the rs8179181 variant in the *TGFB1* gene (OR 0.53, p = 0.0316) and for the rs11556089 variant in the *SMAD3* gene (OR 0.35, p = 0.0416; **Table 4**). In the replication phase on an independent sample, rs8179181 (OR 0.44, p = 0.0073) but not rs11556089 (OR 0.63, p = 0.0535) was again associated with PD. However, joint analysis also showed a weak association of the *SMAD3* rs11556089 variant (OR 0.49, 95% CI 0.26–0.93, p = 0.0494) and further confirmed the association of rs8179181 in the *TGFB1* gene with PD (OR 0.48, 95% CI 0.32–0.73, p = 0.00062).

**Table 4.**
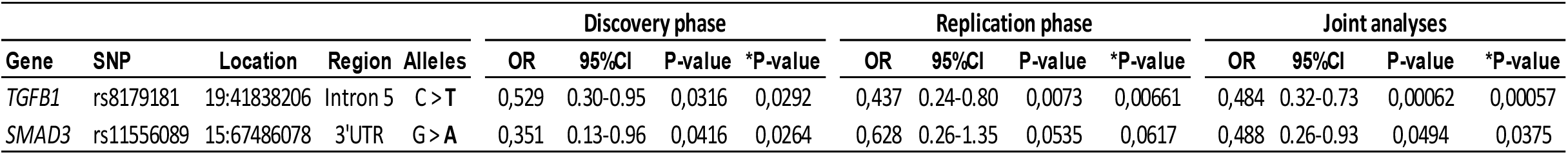
Association of SNPs with PD in the discovery and replication phases, and in the joint analysis. The alleles refer to the major/minor alleles on the forward strand, with the protective allele identified in bold. Association by logistic regression adjusted for age and sex covariates: OR, odds ratio; CI, confidence interval. *P-value adjusted by permutation testing.

We first applied a conservative Bonferroni approach to correct for multiple comparisons and obtained a p-value of 0.1012 for rs8179181. However, this method assumes the independence of all tests. Since many of these markers are likely to be in linkage disequilibrium, we further applied permutation testing as a more robust method for multiple comparisons and considered the method of choice for accurate correction in candidate-gene association studies [34-36]. The permutation testing adjusted the p-value to 0.00057 for rs8179181 and 0.0375 for rs11556089, showing a significant association of rs8179181 with PD and a weaker effect of rs11556089. No differences between the expected and observed heterozygosity were evident for either rs8179181 or rs11556089, nor were there any deviations from HWE (data not shown). None of these SNPs had previously been associated with PD, and a protective effect of the minor allele was evident for both SNPs (rs8179181: MAF = 0.317 and 0.192 in controls and PD patients, respectively; rs11556089: MAF= 0.112 and 0.064 in controls and PD patients, respectively).

### Haplotype-association analysis

Seven distinct haplotype blocks were detected, which corresponded to the 3’UTR of *SMAD3, SMAD2*, and *TGFB2*, or the downstream *TGFB1* gene (**Table 5**). Similar haplotype frequencies were obtained, and the adjusted logistic regression analysis did not identify significant differences between the controls and patients with PD (data not shown).

**Table 5.**
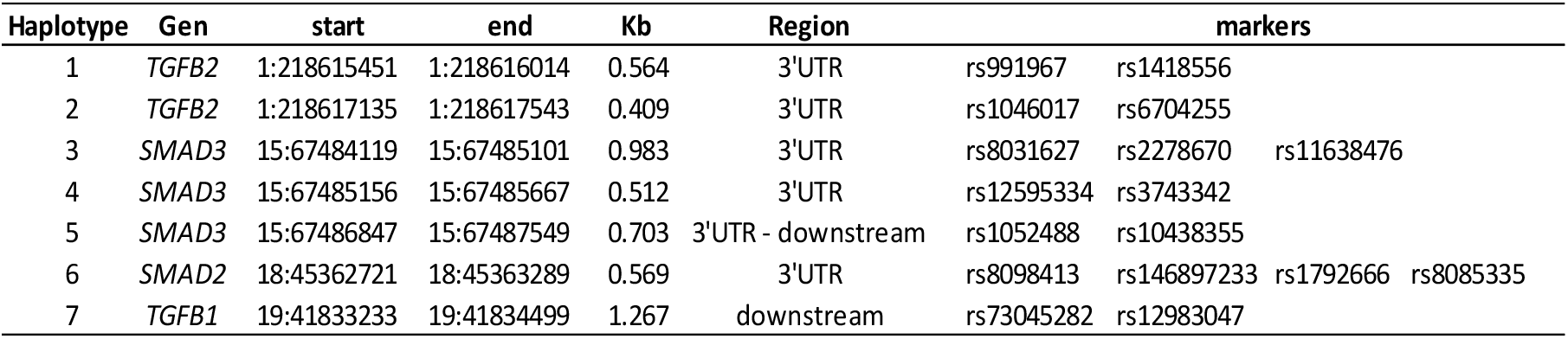
Haplotype block estimation. Start/end position of the block, chromosome:bp units; Kb, distance spanned by the blocks in kilobases; markers, list of SNPs in each block.

## DISCUSSION

Here, we present the results of two independent candidate gene studies and a joint analysis using NGS to investigate the association between idiopathic PD and common variants of genes involved in the TGF-β signaling pathway (*TGFB1, TGFB2, SMAD2, SMAD3, TGFBRI, and TGFBRII*). The results suggest that the rs8179181 variant of the *TGFB1* gene is associated with PD, conferring protection against this disorder. This association was evident in both the discovery phase and in an independent exact replication phase, as well as in the joint analysis, suggesting *TGFB1* as a novel PD susceptibility locus. In fact, rs8179181 was detected as an AIM with the protective minor allele being present at approximately 25–30% in IBS and EUR Caucasian populations, but at only 0.004–0.1% of the AFR, Asian, or AMR populations.

Through several GWAS, 90 independent risk signals have been identified at different loci that are associated with a modest risk of idiopathic PD. GWAS is a hypothesis-free method used to examine common variations associated with a disorder. However, it is estimated that the GWAS loci identified to date explain only a fraction of the genetic component of idiopathic PD [23, 24]. Indeed, a challenge will be to fully characterize and obtain sufficient functional evidence for all variants identified by GWAS; in this sense, other methods such as fine-mapping using NGS of candidate genes can complement this approach. To date, no associations between TGF-β genes and PD have been reported in GWAS, which may be due to the different study settings and populations analyzed. Notably, the results of this study are supported by previous functional studies in mouse models indicating an association between deficient TGF-β signaling and parkinsonism [16-22]. Thus, our approach, based on this prior functional evidence and on the fine mapping of the genes involved in this process, goes one step further than hypothesis-free and tagged SNP methods such as GWAS [25, 26, 39].

A genetic association study based on haplotype tagging showed that the rs6658835 SNP in the 5′ region of *TGFB2* is associated with susceptibility to PD, although with borderline significance [27]. Unfortunately, our approach did not detect this SNP, as it is located in the first intron of the gene, which was not analyzed. Several studies have indicated that the rs1800470 SNP of *TGFB1* is associated with vascular-related dementia, AD, and cognitive alterations [40-44]. This variant is located at codon +10 (T/C) in the signal peptide of the protein; however, in our study, it did not appear to be associated with PD (OR 1.03, 95% CI 0.73–1.46, p = 0.8681).

The rs11556089 *SMAD3* variant, which is weakly associated with PD, is located in the 3’ UTR of the gene, in exon 9, and is conserved in non-human primates. The location of this variant suggests that it may alter the efficiency of mRNA translation, its distribution, its protein-protein interactions, or putative microRNA-binding sites [45]. The rs8179181 *TGFB1* variant (c.861-20C>T) is located in the fifth intron, 20 bp from exon 6 (**Figure 1**). As this locus is highly conserved in different species, it may have a relevant effect on TGF-β signaling. Indeed, it can affect a branch point, which is usually located 14–40 nucleotides upstream of the 3’ end of an intron [46]. Branch-point substitutions can influence gene splicing through exon skipping or cryptic splice site activation. Moreover, if the variant were to impede the removal of the intron, an incorrect protein might be produced with extra “junk” information [47].

**Figure 1.**
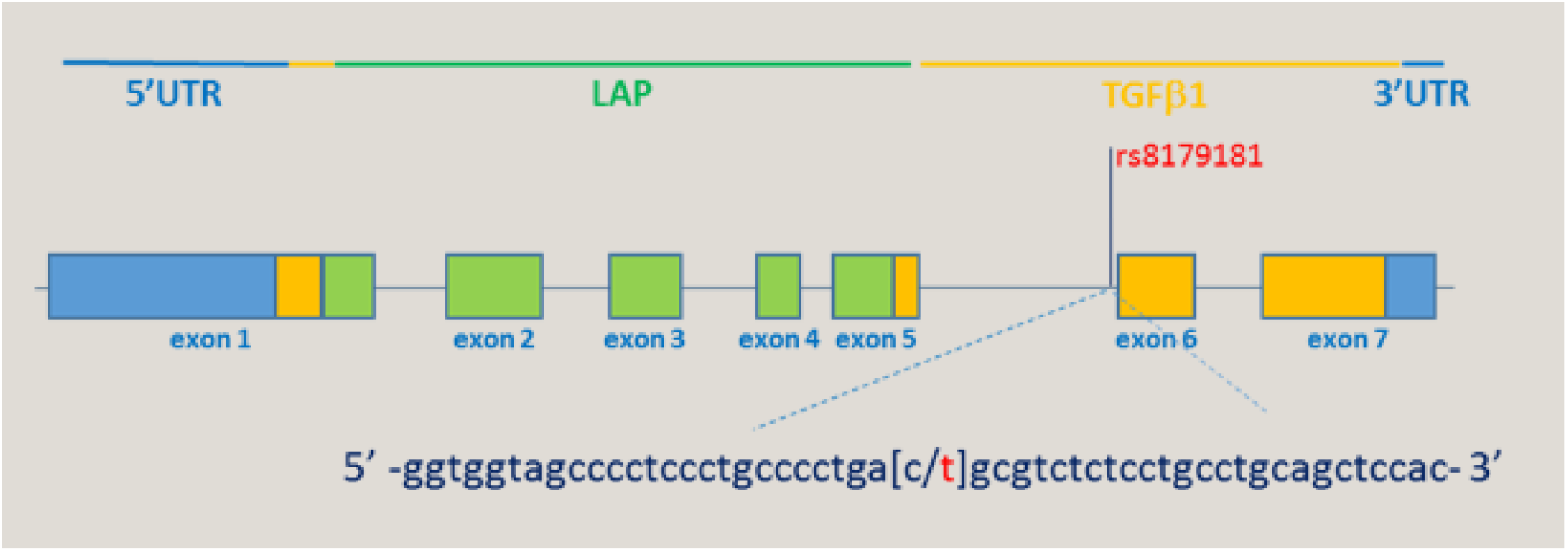
Schematic structure of the *TGFB1* gene displaying the relationships between the exons and the functional domains. The rs8179181 polymorphism that was seen to be associated with PD could affect a branch point and gene splicing: LAP (green); latent associated protein; TGFβ1 (yellow), mature protein.

The functional significance of this variant remains unclear, although TGF-β1 is known to influence the balance between bone formation and resorption [48] and the rs8179181 variant has previously been associated with osteoporosis and altered bone mass [49, 50]. Interestingly, PD patients have more osteoporotic fractures than healthy subjects [51], and the association of the rs8179181 SNP with PD might be related to this non-motor symptom of the disorder. TGF-β1 is also a molecule central to adaptive immunity, either inhibiting or enhancing T cell proliferation or disrupting the survival or accumulation of these cells at specific tissue sites. TGF-β also plays a central role in the differentiation of T cells, mainly in the regulation of effector and regulatory CD4(+) T cells (Th1, Th2, Treg, Th17, and Th9 cells), controlling both anti-inflammatory and pro-inflammatory T cell responses [52]. Emerging evidence suggests that the adaptive immune system is involved in the pathogenesis of PD. Patients have increased levels of infiltrating T cells, which may be autoreactive against altered self-aggregated α-synuclein, although T-cell nigral infiltration precedes synucleinopathy in the early stages of the disease [53-54]. The role of TGF-β1 in the adaptive immune system of PD patients remains unknown, although Smad3-deficient mice develop α-synuclein/ubiquitin inclusions in different brain areas, including the SN, striatum, and motor cortices, with a core/halo morphology resembling that of human LBs [17]. Thus, it would be interesting to evaluate whether T cells participate in α-synuclein aggregation in the brain of Smad3 deficient mice. Nevertheless, we cannot rule out the possibility that TGF-β1/Smad3 signaling directly regulates the survival of dopaminergic neurons without the participation of immune cells.

## CONCLUSIONS

In summary, our data show that the rs8179181 variant of the TGFB1 gene is associated with PD, which was reproducible in independent samples. Defining the events affected by this variant could provide key insights into the pathogenesis of PD, leading to the development of novel and improved diagnostic and therapeutic approaches.

## Data Availability

All data produced in the present study are available upon reasonable request to the authors

## ABBREVIATIONS

AD: Alzheimer’s disease
AFR: African population
AIMs: ancestry informative markers
AMR: Ad-Mixed American population
bp: base pair
CI: confidence interval
EAS: East Asian population
EUR: European Caucasian population
GWAS: genome-wide association study
HWE: Hardy-Weinberg equilibrium
IBS: Spanish population
LB: Lewy body
MAF: minor allele frequency
NGS: next-generation sequencing
OR: odds ratio
PD: Parkinson’s disease
SAS: South Asian population
SN: substantia nigra
SNP: single nucleotide polymorphism
SNV: single nucleotide variation
TGF-β: transforming growth factor-beta
UTR: untranslated region

## ACKNOWLEDGMENTS

This work was supported by the Instituto de Salud Carlos III - ISCIII (PI16/00186; Plan Estatal de I+D+I 2013-2016) and was co-financed by the European Development Regional Fund “A way to achieve Europe” (ERDF). The funders played no role in the study design; in the collection, analysis and interpretation of data; in the writing of the report; and in the decision to submit the article for publication.

## REFERENCES

[1] E.R. Dorsey, B.R. Bloem, The Parkinson pandemic-A call to action, JAMA Neurol. 75 (2018) 9–10.

[2] A. Schrag, Z. Anastasiou, G. Ambler, A. Noyce, K. Walters, Predicting diagnosis of Parkinson’s disease: A risk algorithm based on primary care presentations, Mov. Disord. 34 (2019) 480–486.

[3] D.W. Dickson, Neuropathology of Parkinson disease, Parkinsonism Relat. Disord. 46 Suppl 1 (2018) S30–S33.

[4] R. Giráldez-Pérez, M. Antolín-Vallespín, M. Muñoz, A. Sánchez-Capelo, Models of alpha-synuclein aggregation in Parkinson’s disease, Acta Neuropathol. Commun. 2 (2014) 176.

[5] M.D. Muñoz, N. de la Fuente, A. Sánchez-Capelo, TGF-beta/Smad3 signalling modulates GABA neurotransmission: Implications in Parkinson’s disease, Int. J. Mol. Sci. 21 (2020) 590.

[6] R. Derynck, E.H. Budi, Specificity, versatility, and control of TGF-beta family signaling, Sci. Signal. 12 (2019).

[7] A. Sánchez-Capelo, Dual role for TGF-beta1 in apoptosis, Cytokine Growth Factor Rev. 16 (2005) 15–34.

[8] M. Mogi, M. Harada, T. Kondo, et al., Transforming growth factor-beta 1 levels are elevated in the striatum and in ventricular cerebrospinal fluid in Parkinson’s disease, Neurosci. Lett. 193 (1995) 129–132.

[9] M.P. Vawter, O. Dillon-Carter, W.W. Tourtellotte, P. Carvey, W.J. Freed, TGFbeta1 and TGFbeta2 concentrations are elevated in Parkinson’s disease in ventricular cerebrospinal fluid, Exp. Neurol. 142 (1996) 313–322.

[10] E.A. van der Wal, F. Gómez-Pinilla, C.W. Cotman, Transforming growth factor-beta 1 is in plaques in Alzheimer and Down pathologies, Neuroreport 4 (1993) 69–72.

[11] H. Zetterberg, N. Andreasen, K. Blennow, Increased cerebrospinal fluid levels of transforming growth factor-beta1 in Alzheimer’s disease, Neurosci. Lett. 367 (2004) 194–196.

[12] A. Sánchez-Capelo, P. Colin, B. Guibert, N.F. Biguet, J. Mallet, Transforming growth factor beta1 overexpression in the nigrostriatal system increases the dopaminergic deficit of MPTP mice, Mol. Cell. Neurosci. 23 (2003) 614–625.

[13] A. Sánchez-Capelo, O. Corti, J. Mallet, Adenovirus-mediated over-expression of TGFbeta1 in the striatum decreases dopaminergic cell survival in embryonic nigral grafts, Neuroreport 10 (1999) 2169–2173.

[14] U. Ueberham, E. Ueberham, M.K. Brückner, et al., Inducible neuronal expression of transgenic TGF-beta1 in vivo: dissection of short-term and long-term effects, Eur. J. Neurosci. 22 (2005) 50–64.

[15] A. Martínez-Canabal, A.L. Wheeler, D. Sarkis, et al., Chronic over-expression of TGFβ1 alters hippocampal structure and causes learning deficits, Hippocampus 23 (2013) 1198–1211.

[16] Z.B. Andrews, H. Zhao, T. Frugier, et al., Transforming growth factor beta2 haploinsufficient mice develop age-related nigrostriatal dopamine deficits, Neurobiol. Dis. 21 (2006) 568–575.

[17] S. Tapia-González, R.M. Giráldez-Pérez, M.I. Cuartero, et al., Dopamine and alpha-synuclein dysfunction in Smad3 null mice, Mol. Neurodeg. 6 (2011) 72.

[18] I. Tesseur, A. Nguyen, B. Chang, et al., Deficiency in neuronal TGF-beta signaling leads to nigrostriatal degeneration and activation of TGF-beta signaling protects against MPTP neurotoxicity in mice, J. Neurosci. 37 (2017) 4584–4592.

[19] S. Villapol, Y. Wang, M. Adams, A.J. Symes, Smad3 deficiency increases cortical and hippocampal neuronal loss following traumatic brain injury, Exp. Neurol. 250 (2013) 353–365.

[20] M. Muñoz, M. Antolín-Vallespín, S. Tapia-González, A. Sánchez-Capelo, Smad3 deficiency inhibits dentate gyrus LTP by enhancing GABAA neurotransmission, J. Neurochem. 137 (2016) 190–199.

[21] S. Tapia-González, M.D. Muñoz, M.I. Cuartero, A. Sánchez-Capelo, Smad3 is required for the survival of proliferative intermediate progenitor cells in the dentate gyrus of adult mice, Cell Commun. Signal. 11 (2013) 93.

[22] S.X. Luo, L. Timbang, J.I. Kim, et al., TGF-beta signaling in dopaminergic neurons regulates dendritic growth, excitatory-inhibitory synaptic balance, and reversal learning, Cell Rep. 17 (2016) 3233–3245.

[23] C. Blauwendraat, M.A. Nalls, A.B. Singleton, The genetic architecture of Parkinson’s disease, The Lancet. Neurology 19 (2020) 170–178.

[24] M.F. Keller, M. Saad, J. Bras, et al., Using genome-wide complex trait analysis to quantify ‘missing heritability’ in Parkinson’s disease, Hum. Mol. Genet. 21 (2012) 4996–5009.

[25] S.E. Pierce, A. Booms, J. Prahl, et al., Post-GWAS knowledge gap: the how, where, and when, NPJ Parkinsons Dis. 6 (2020) 23.

[26] D.C. Koboldt, K.M. Steinberg, D.E. Larson, R.K. Wilson, E.R. Mardis, The next-generation sequencing revolution and its impact on genomics, Cell 155 (2013) 27–38.

[27] A. Goris, C.H. Williams-Gray, T. Foltynie, et al., Investigation of TGFB2 as a candidate gene in multiple sclerosis and Parkinson’s disease, J. Neurol. 254 (2007) 846–848.

[28] A.D. Skol, L.J. Scott, G.R. Abecasis, M. Boehnke, Joint analysis is more efficient than replication-based analysis for two-stage genome-wide association studies, Nat. Genet. 38 (2006) 209–213.

[29] A. Albanese, Diagnostic criteria for Parkinson’s disease, Neurol. Sci. 24 Suppl 1 (2003) S23–26.

[30] A.J. Hughes, S.E. Daniel, L. Kilford, A.J. Lees, Accuracy of clinical diagnosis of idiopathic Parkinson’s disease: a clinico-pathological study of 100 cases, J. Neurol. Neurosurg. Psychiatry 55 (1992) 181–184.

[31] C.B. Breckenridge, C. Berry, E.T. Chang, et al., Association between Parkinson’s disease and cigarette smoking, rural living, well-water consumption, farming and pesticide use: Systematic review and meta-analysis, PloS One 11 (2016) e0151841.

[32] C.A. Anderson, F.H. Pettersson, G.M. Clarke, et al., Data quality control in genetic case-control association studies, Nat. Protoc. 5 (2010) 1564–1573.

[33] T.L. Edwards, X. Gao, Methods for detecting and correcting for population stratification, Curr. Protoc. Hum. Genet. Chapter 1, (2012) 22, 21–14.

[34] N.-N.W.G.o.R.i.A. Studies, S.J. Chanock, T. Manolio, et al., Replicating genotype-phenotype associations, Nature 447 (2007) 655–660.

[35] G.M. Clarke, C.A. Anderson, F.H. Pettersson, et al., Basic statistical analysis in genetic case-control studies, Nat. Protoc. 6 (2011) 121–133.

[36] P.H. Westfall, S.S. Young, Resampling-based multiple testing: Examples and methods for P-value adjustment, John Wiley & Sons, New York, 1993.

[37] M.A. Quail, M. Smith, P. Coupland, et al., A tale of three next generation sequencing platforms: comparison of Ion Torrent, Pacific Biosciences and Illumina MiSeq sequencers, BMC Genomics 13 (2012) 341.

[38] E.A. Worthey, Analysis and annotation of whole-genome or whole-exome sequencing-derived variants for clinical diagnosis, Curr. Protoc. Hum. Genet. 79 (2013) Unit 9 24.

[39] R.V. Broekema, O.B. Bakker, I.H. Jonkers, A practical view of fine-mapping and gene prioritization in the post-genome-wide association era, Open Biol. 10 (2020) 190221.

[40] B. Arosio, L. Bergamaschini, L. Galimberti, et al., +10 T/C polymorphisms in the gene of transforming growth factor-beta1 are associated with neurodegeneration and its clinical evolution, Mech. Ageing Dev. 128 (2007) 553–557.

[41] F. Caraci, P. Bosco, M. Signorelli, et al., The CC genotype of transforming growth factor-beta1 increases the risk of late-onset Alzheimer’s disease and is associated with AD-related depression, Eur. Neuropsychopharmacol. 22 (2012) 281–289.

[42] D. Frydecka, B. Misiak, E. Pawlak-Adamska, et al., Sex differences in TGFB-beta signaling with respect to age of onset and cognitive functioning in schizophrenia, Neuropsychiatr. Dis. Treat. 11 (2015) 575–584.

[43] Q. Yang, E.Y. Wang, H.W. Jia, Y.P. Wang, Association between polymorphisms in transforming growth factor-beta1 and sporadic Alzheimer’s disease in a Chinese population, Int. J. Neurosci. 126 (2016) 979–984.

[44] R. Peila, B. Yucesoy, L.R. White, et al., A TGF-beta1 polymorphism association with dementia and neuropathologies: the HAAS, Neurobiol. Aging 28 (2007) 1367–1373.

[45] C. Mayr, Regulation by 3’-Untranslated Regions, Annu. Rev. Genet. 51 (2017) 171–194.

[46] M.E. Wilkinson, C. Charenton, K. Nagai, RNA Splicing by the Spliceosome, Annu. Rev. Biochem. 89 (2020) 359–388.

[47] J. Kralovicova, H. Lei, I. Vorechovsky, Phenotypic consequences of branch point substitutions, Hum. Mutat. 27 (2006) 803–813.

[48] Y. Tang, X. Wu, W. Lei, et al., TGF-beta1-induced migration of bone mesenchymal stem cells couples bone resorption with formation, Nat. Med. 15 (2009) 757–765.

[49] R.W. Keen, H. Snieder, H. Molloy, et al., Evidence of association and linkage disequilibrium between a novel polymorphism in the transforming growth factor beta 1 gene and hip bone mineral density: a study of female twins, Rheumatol. 40 (2001) 48–54.

[50] B.L. Langdahl, M. Carstens, L. Stenkjaer, E.F. Eriksen, Polymorphisms in the transforming growth factor beta 1 gene and osteoporosis, Bone 32 (2003) 297–310.

[51] V. Metta, T.C. Sánchez, C. Padmakumar, Osteoporosis: A Hidden Nonmotor Face of Parkinson’s Disease, Int. Rev. Neurobiol. 134 (2017) 877–890.

[52] M.A. Travis, D. Sheppard, TGF-beta activation and function in immunity, Annu. Rev. Immunol. 32 (2014) 51–82.

[53] J. Galiano-Landeira, A. Torra, M. Vila, J. Bove, CD8 T cell nigral infiltration precedes synucleinopathy in early stages of Parkinson’s disease, Brain 143 (2020) 3717–3733.

[54] D. Sulzer, R.N. Alcalay, F. Garretti, et al., T cells from patients with Parkinson’s disease recognize alpha-synuclein peptides, Nature 546 (2017) 656–661.

