## Supplementary Material for "*TGFB1* rs8179181 polymorphism is reproducibly associated with Parkinson’s disease in a Spanish population"

**Supplementary Material S1**. **Coding and regulatory regions targeted for resequencing**. Start/end position in base pairs (bp), according to the NCBI build GRCh37.

| Gene | Region | Chro. | Start | End | Num. Amplicons | Size (bases) |
|---|---|---|---|---|---|---|
| TGFB1 | 5'UTR/coding | chr19 | 41836806 | 41837120 | 2 | 314 |
| TGFB1 | coding/splicing | chr19 | 41838027 | 41838191 | 2 | 164 |
| TGFB1 | coding/splicing | chr19 | 41847782 | 41847940 | 1 | 158 |
| TGFB1 | coding/splicing | chr19 | 41848069 | 41848157 | 1 | 88 |
| TGFB1 | coding/splicing | chr19 | 41850646 | 41850774 | 1 | 128 |
| TGFB1 | coding/splicing | chr19 | 41854194 | 41854365 | 2 | 171 |
| TGFB1 | coding/3'UTR | chr19 | 41858589 | 41859836 | 4 | 1247 |
| TGFB2 | 5'UTR/coding | chr1 | 218518670 | 218520394 | 10 | 1724 |
| TGFB2 | coding/splicing | chr1 | 218536670 | 218536764 | 1 | 94 |
| TGFB2 | coding/splicing | chr1 | 218578505 | 218578679 | 2 | 174 |
| TGFB2 | coding/splicing | chr1 | 218607418 | 218607561 | 1 | 143 |
| TGFB2 | coding/splicing | chr1 | 218607674 | 218607795 | 1 | 121 |
| TGFB2 | coding/splicing | chr1 | 218609306 | 218609494 | 1 | 188 |
| TGFB2 | coding/splicing | chr1 | 218610679 | 218610843 | 1 | 164 |
| TGFB2 | coding/3'UTR | chr1 | 218614540 | 218617966 | 18 | 3426 |
| SMAD3 | 5'UTR/coding | chr15 | 67358189 | 67358703 | 3 | 514 |
| SMAD3 | 5'UTR | chr15 | 67418048 | 67418327 | 3 | 279 |
| SMAD3 | 5'UTR/coding | chr15 | 67430353 | 67430443 | 1 | 90 |
| SMAD3 | coding/splicing | chr15 | 67457227 | 67457431 | 2 | 204 |
| SMAD3 | coding/splicing | chr15 | 67457585 | 67457727 | 2 | 142 |
| SMAD3 | 5'UTR | chr15 | 67458487 | 67458536 | 1 | 49 |
| SMAD3 | coding/splicing | chr15 | 67459111 | 67459196 | 1 | 85 |
| SMAD3 | coding/splicing | chr15 | 67462886 | 67462947 | 1 | 61 |
| SMAD3 | coding/splicing | chr15 | 67473573 | 67473796 | 2 | 223 |
| SMAD3 | coding/splicing | chr15 | 67477059 | 67477207 | 1 | 148 |
| SMAD3 | coding/splicing | chr15 | 67479697 | 67479852 | 1 | 155 |
| SMAD3 | coding/3'UTR | chr15 | 67482745 | 67487538 | 25 | 4793 |
| SMAD2 | 5'UTR/coding | chr18 | 45359460 | 45368326 | 46 | 8866 |
| SMAD2 | coding/splicing | chr18 | 45371705 | 45371860 | 2 | 155 |
| SMAD2 | coding/splicing | chr18 | 45372028 | 45372176 | 1 | 148 |
| SMAD2 | coding/splicing | chr18 | 45374840 | 45375063 | 2 | 223 |
| SMAD2 | coding/splicing | chr18 | 45377639 | 45377703 | 1 | 64 |
| SMAD2 | coding/splicing | chr18 | 45391424 | 45391509 | 1 | 85 |
| SMAD2 | coding/splicing | chr18 | 45394688 | 45394833 | 1 | 145 |
| SMAD2 | coding/splicing | chr18 | 45395608 | 45395812 | 2 | 204 |
| SMAD2 | coding/splicing | chr18 | 45396840 | 45396940 | 1 | 100 |
| SMAD2 | coding/3'UTR | chr18 | 45422886 | 45423185 | 2 | 299 |
| SMAD2 | 3'UTR | chr18 | 45456726 | 45456931 | 1 | 205 |
| SMAD2 | 3'UTR | chr18 | 45457165 | 45457517 | 1 | 352 |
| SMAD2 | 3'UTR | chr18 | 45457165 | 45457520 | 1 | 355 |
| TGFBR1 | 5'UTR/coding | chr9 | 101867406 | 101867589 | 0 | 183 |
| TGFBR1 | coding/splicing | chr9 | 101891131 | 101891387 | 2 | 256 |
| TGFBR1 | coding/splicing | chr9 | 101894785 | 101895026 | 2 | 241 |
| TGFBR1 | coding/splicing | chr9 | 101900135 | 101900376 | 2 | 241 |
| TGFBR1 | coding/splicing | chr9 | 101904812 | 101904990 | 1 | 178 |
| TGFBR1 | coding/splicing | chr9 | 101907008 | 101907175 | 2 | 167 |
| TGFBR1 | coding/splicing | chr9 | 101908761 | 101908896 | 1 | 135 |
| TGFBR1 | coding/splicing | chr9 | 101909930 | 101910071 | 1 | 141 |
| TGFBR1 | coding/3'UTR | chr9 | 101911456 | 101916478 | 28 | 5022 |
| TGFBR2 | 5'UTR/coding | chr3 | 30647988 | 30648474 | 3 | 486 |
| TGFBR2 | coding/splicing | chr3 | 30664685 | 30664770 | 1 | 85 |
| TGFBR2 | coding/splicing | chr3 | 30686233 | 30686412 | 2 | 179 |
| TGFBR2 | coding/splicing | chr3 | 30691756 | 30691957 | 1 | 201 |
| TGFBR2 | coding/splicing | chr3 | 30713124 | 30713934 | 5 | 810 |
| TGFBR2 | coding/splicing | chr3 | 30715591 | 30715743 | 1 | 152 |
| TGFBR2 | coding/splicing | chr3 | 30729870 | 30730008 | 1 | 138 |
| TGFBR2 | coding/3'UTR | chr3 | 30732906 | 30735638 | 15 | 2732 |

**Supplementary material S2**. **Description of the AmpliSeq™ primer system.** [1]Chromosome positions according to the NCBI build GRCh37.

| Gene | Primer sequence (5'->3') Forward | Reverse | Amplicon postion[1] | Pool |
|---|---|---|---|---|
| TGFB1 | GCTGAGGTCCTCAGGGAGAAG | CGCGCATCCTAGACCCTTT | chr19:41859069-41859257 | 2 |
| TGFB1 | GTCAGCACCAGTAGCCACA | CCGTGGGATACTGAGACACC | chr19:41858884-41859142 | 1 |
| TGFB1 | CTTCACCAGCTCCATGTCGAT | CGAGGCCCTCCTACCTTTTG | chr19:41858823-41859062 | 2 |
| TGFB1 | ACCGTTGTGGGTTTCCACC | GGACTATCCACCTGCAAGACT | chr19:41858592-41858865 | 1 |
| TGFB1 | TTCTCGGAGCTCTGATGTGTTG | GTCAGCTCCAAAACTCCTAGAGTTG | chr19:41854289-41854449 | 2 |
| TGFB1 | GGACCATCTAGGTGGACCTTGT | GCAGAGTACACACAGCATATATATGTTCTT | chr19:41854091-41854341 | 1 |
| TGFB1 | CCAGGTCTCAGCACTTTCACAC | GTCTCTCTCCTGCCTTCATCATC | chr19:41850528-41850798 | 2 |
| TGFB1 | TACACACACACTCTCAGAGGGATAG | GGTTTGCTCCTTCCTTCCTCTTC | chr19:41847969-41848233 | 1 |
| TGFB1 | GAGCACCTGGTCAGCAGAT | ATCCCTCTGAGAGTGTGTGTGTA | chr19:41847717-41847992 | 2 |
| TGFB1 | TTGCGGAAGTCAATGTACAGCT | CAAGGGAGACCCAGATGGAGATA | chr19:41838135-41838316 | 1 |
| TGFB1 | GCCAACTCACCTCTCTGACTTTA | CTGCAGCTCCACGGAGAAGAAC | chr19:41837934-41838192 | 2 |
| TGFB1 | CGCCCAATGACACAGAGATC | CTGCCCATCGTGTACTACGT | chr19:41836793-41837040 | 1 |
| TGFB1 | GGAACGTCAGGGATGGAGAC | GCTGTCCAACATGATCGTGC | chr19:41836763-41836996 | 2 |
| TGFB1 | GGCATACAGTGGCACAGTCAG | TGAAGAAGACAATAAAAGATAGTAGTTCAGGC | chr19:41836200-41836468 | 1 |
| TGFB1 | GAGAAAGAGGAAGCAGGAGAGAATTC | AAAAAGAACTTGCACTCAAGGAAGA | chr19:41834975-41835218 | 2 |
| TGFB1 | CTCATGCCCACAGGCAAAGGAAC | CATCTGCTATGGTCCTGCTTGT | chr19:41834864-41835023 | 1 |
| TGFB1 | CTCTATACCTATCCATTTGTTGGCATGT | GGAGGAAGGACAGTATGAGTGCT | chr19:41834686-41834913 | 2 |
| TGFB1 | GTACACGCACCACATCCTCATA | AGGCAGGAGTTCACTATGCAAG | chr19:41834591-41834737 | 1 |
| TGFB1 | TTTACGTGATGGGCTGACACTT | ACACAGTGACTCGGCTGTAATC | chr19:41834398-41834645 | 2 |
| TGFB1 | TGTTGAGGGATCCATTTCCTGTC | AGCCCACATATACACAGATACAAATAGAGA | chr19:41834198-41834450 | 1 |
| TGFB1 | GGAGGGTGTGGGATGTCAGA | ACGCATCACGGCATCAGA | chr19:41834029-41834244 | 2 |
| TGFB1 | GACCACACCGCGAATGTG | GGTCGGTCTTACCCACTCAG | chr19:41833818-41834069 | 1 |
| TGFB1 | ACGGAGCTAACACGTGCTG | GGGAGATTGGTGGGTGAGAGAA | chr19:41833137-41833363 | 2 |
| TGFB1 | AATGCACAAATTTACACTCAGAAGAACA | GCTGTTTGGTATGGATGGTCTTGT | chr19:41832940-41833193 | 1 |
| TGFB2 | CAGACACGTGGTTCAGAGAGAA | TTTGTTCCTGGATGACTCCCTAGA | chr1:218518624-218518872 | 1 |
| TGFB2 | GGGATGGAGAATATTAGCCTGACG | CCCAACGTGGAATTGCTCG | chr1:218518823-218518969 | 2 |
| TGFB2 | GCAAGTGCCTACCTACCCTAAG | CTCCTCCTTCTCTTGCTCCAAAC | chr1:218518928-218519153 | 1 |
| TGFB2 | AAGGGCTGCCGTTGTGAT | GCTGACCCGCTTGGTTACT | chr1:218519060-218519232 | 2 |
| TGFB2 | CACACACGTGTGCGCTTC | GCGGATCTGAACTCGGCT | chr1:218519428-218519661 | 1 |
| TGFB2 | GACCTTTTCATCTCTTCCCTTTTGG | CCTGACTTTGGCGAGTAAGAAGAAA | chr1:218519611-218519797 | 2 |
| TGFB2 | CCCATCTCATTGCTCCAAGAATTTT | GCGAAACTTTTGCAAACAATCTAGTCA | chr1:218519747-218519923 | 1 |
| TGFB2 | AGGATACGTTTTTCTGTTGGGCAT | TGCGCATGAACTGGTCCATATC | chr1:218519872-218520146 | 2 |
| TGFB2 | TGTCTACCTGCAGCACACTC | GCAAGTCCCTGGTGCTGTT | chr1:218520104-218520275 | 1 |
| TGFB2 | CCCGGAGGTGATTTCCATCTAC | CGGGCAGAGCTAAACCTCAG | chr1:45395677-218520436 | 2 |
| TGFB2 | GATAAGAACATTCAATGAGTCCCTCTTCT | CCCACAGGAGACAAACACTAAAGATG | chr1:218536567-218536836 | 1 |
| TGFB2 | TAATGGTATTAAACTGGCCGTTGGAA | GAGACGTCAAATCGAACAATTCTGAAG | chr1:218578372-218578568 | 2 |
| TGFB2 | CGCCCACTTTCTACAGACCCTA | GAGAGAGAGAGTCTGTCTAAAACAACAAC | chr1:218578519-218578737 | 1 |
| TGFB2 | TCATGCTGTCAGAATGCCAACT | TGTGTCTATACCCTTAAATCTGCAATCAAT | chr1:218607369-218607644 | 2 |
| TGFB2 | TGATTGCAGATTTAAGGGTATAGACACAC | CCAGTCTCATTTTCACATGCAAACT | chr1:218607616-218607887 | 1 |
| TGFB2 | GGTGAAGCTAAATGTTTATTACCCAAATGC | GCAGTTTAAGAGTTAACAGATGCAATTACT | chr1:218609270-218609543 | 2 |
| TGFB2 | GTGAGGGTGGTGAATCAGCTTT | TATCAGCAGGTGCACATGGTAAC | chr1:218610626-218610897 | 1 |
| TGFB2 | AAAAACGAATTGCGTTCATTTTCCG | CATTGTCATTTTGGTCTTGCCACTTT | chr1:218614480-218614740 | 2 |
| TGFB2 | TTGCAAATGCAGCTAAAATTCTTGGA | TTTGCTTCTTGTCTCTCTCACTTACAAA | chr1:218614688-218614963 | 1 |
| TGFB2 | TTGAAGGCCTTATTCTACATTTCACCTAC | AAGGGTGCCTATTGCATAGCAAT | chr1:218614906-218615141 | 2 |
| TGFB2 | GCGCCTCACTTGATTTTTCTGT | CAGACTTTCTCGGTCATATAATAACTCACT | chr1:218615096-218615369 | 1 |
| TGFB2 | GAGGTCATATTAATGGGATGAAAACCCA | ACCAGTACATGTCAAAACATTTCAACATTT | chr1:218615311-218615577 | 2 |
| TGFB2 | GGTTCTTTGTCAGTTTAGTAAACCAGTG | AAAGAGATCTGCAGATGTGTTTTCAATT | chr1:218615519-218615719 | 1 |
| TGFB2 | ATTATTAAAATAGATGGATATAGAAGCCAGCAT | TACTGGCCAGCTAAAAGAAACATACATT | chr1:218615658-218615883 | 2 |
| TGFB2 | TGGGCTCTTTTTAATGATCACTCACA | CGAACCCATCTTTTCTGTGAAACAAAAA | chr1:218615829-218616045 | 1 |
| TGFB2 | CCTGGGTCCACTTGTCTTTTCTTT | AAACCAAGGTAGAGAAACTTTTCTTAACTC | chr1:218615993-218616241 | 2 |
| TGFB2 | GCAAGGATTTAGGGTTCTAACTAAAACTCA | ATGACTTATGCTGACCTGGACTTTTT | chr1:218616170-218616403 | 1 |
| TGFB2 | CAGCATAAGTCATTTTGTGTATTTCACTGA | CTAAATAAGCGACAAGTGATAAACTGACAT | chr1:218616390-218616639 | 2 |
| TGFB2 | TGCAGAAGAGGTATCCTCATGCT | TATTATGGCATCCCATCTCTTGCTTTT | chr1:218616572-218616834 | 1 |
| TGFB2 | GAATTGGATTTGCAAGTTTGAAAACTGG | CAGTGCATTCATGAGGAAAGACAAAATAAT | chr1:218616779-218617010 | 2 |
| TGFB2 | CCTCTCAGATAGGATGACATTTTGTTTTGT | CGATCAAATTGTACTGCTGATTAACACAAA | chr1:218616950-218617209 | 1 |
| TGFB2 | GTATGCAAGTGGGCAGCAATTATT | ATGACTTTTCCAGCTTGAGATAGTCTTAT | chr1:218617155-218617425 | 2 |
| TGFB2 | GAGTCTAGAAAACACCTGTGATGCA | GGTGCAAACATCTCTCTGCATTTTT | chr1:218617371-218617645 | 1 |
| TGFB2 | GAGATTTAGTTTCCATCCAGCTTGACT | AGTGGTACCAGTGAAACTGACAG | chr1:218617583-218617831 | 2 |

| | | | | |
|---|---|---|---|---|
| TGFB2 | GAGATTTAGTTTCCATCCAGCTTGACT | AGTGGTACCAGTGAAACTGACAG | chr1:218617583-218617831 | 2 |
| TGFB2 | TGGTTGTTTGGGATCAGCTACTTG | TAGTTCTTTCACTAGACAAAGCAAGACAA | chr1:218617783-218617995 | 1 |
| SMAD3 | CCCAATGTGCCAATTGGTTTGT | GAGCCTCAAGGACATCTGTTGA | chr15:67356951-67357216 | 1 |
| SMAD3 | CCCTCCATTCATTCCTTCAAGAAC | CCCTGGGCTGTTTTTATTCTCTAG | chr15:67357170-67357405 | 2 |
| SMAD3 | CCGGTGTGATCTCACCGA | GGGTTGGACTCGCAGCAA | chr15:67357363-67357569 | 1 |
| SMAD3 | GCGGTGACAGCACTTGGAAA | GCTAGCTTTGATGCGCAGAT | chr15:67357896-67358089 | 2 |
| SMAD3 | CTGCACGCGGATTTGCAT | GCGAGACTCCAAGTGGCAG | chr15:67357921-67358196 | 1 |
| SMAD3 | CGTGCGGAAACCCAAACTT | CGCCCAAACTTCGCCTCA | chr15:67358155-67358390 | 2 |
| SMAD3 | GGCAACTTCGCCGAGAGT | CGTTCTGCTCGCCCTTCTT | chr15:67358354-67358565 | 1 |
| SMAD3 | CGTCCATCCTGCCTTTCACTC | GGGATGGTGATGCACTTGGT | chr15:67358496-67358695 | 2 |
| SMAD3 | TTTTTAAGACAACTGACTCCTCTGAGT | CCACATGTTGCTCTTATGGTCAGT | chr15:67418001-67418171 | 1 |
| SMAD3 | GGGAGCAAACGGCCTGAAATAT | GGTTTCCATGCAGTTGTCCCAT | chr15:67418037-67418288 | 2 |
| SMAD3 | GTTCCCACAGGATGCGACAT | GTTTCTGCTTCTCAGTGACAAATACC | chr15:67418237-67418387 | 1 |
| SMAD3 | GAAAATAGGAGGTACTAGAATTCGGACTTC | CGGAGAAACTCTGACAGATCTCTG | chr15:67430262-67430537 | 2 |
| SMAD3 | CTCTGCAGAAAGCAAGCACAATC | GCATACCTGGTGTCTCTACTCTCTG | chr15:67457170-67457432 | 1 |
| SMAD3 | CTGCGTGAATCCCTACCACTAC | ATGCCAGACCAGCACCAAAGTC | chr15:67457385-67457552 | 2 |
| SMAD3 | GGACAGTTCTACCTCCTGTGTTG | CGTGCCTACCTGGAATATTGCTC | chr15:67457584-67457731 | 1 |
| SMAD3 | GGACGACTACAGCCATTCCATC | CAACAGGGATGCGGTTCTGA | chr15:67457648-67457887 | 2 |
| SMAD3 | GCTGCGTTCCTCTTAGAGCATT | AGTTCAATGACGGCAAAGACAGT | chr15:67458365-67458638 | 1 |
| SMAD3 | GTGTGATGTCTTTGCAAAAGGTGT | CAAATACTGGAGAGAGAGATAGAAGGGA | chr15:67459037-67459311 | 2 |
| SMAD3 | GCTCTCCAGGCCAAGAATCTTT | TCACCGTAACCAGAAACCTGATG | chr15:67462837-67463091 | 1 |
| SMAD3 | GGAAATGGTTTTCCAGAGTGTCCAT | GAAGCCATCCACAGTCATGGAT | chr15:67473485-67473700 | 2 |
| SMAD3 | ATCTCCTACTACGAGCTGAACCA | CCTTCAGAGGCTGTGTGTTCAG | chr15:67473622-67473883 | 1 |
| SMAD3 | ACCCTGTCCAGTCTAACCTGAA | GCAGAAAAGGTGAGAAAGGTGGA | chr15:67477033-67477296 | 2 |
| SMAD3 | GGGTCCAGGACTTGCTTTATCC | CTGATGTAGGCAGCACCCATAA | chr15:67479617-67479874 | 1 |
| SMAD3 | CCCTTTCCCTATTTCTTACAGGAGA | CAACAATGGGTTGAGTAGAGTTCCA | chr15:67482729-67482964 | 2 |
| SMAD3 | CCATGCAGGAGGTGGAGAAAAT | GCCGGTGGTGTAATACTACCTG | chr15:67482917-67483167 | 1 |
| SMAD3 | CTGGTGGCTTAAGTGAGCAGAA | GCTGGGAATGTTCACTGGAAGA | chr15:67483123-67483369 | 2 |
| SMAD3 | CTCTCTGACGCTTGTGACAGT | CCTCCCAATCAGTATGTTCTGAGAG | chr15:67483323-67483497 | 1 |
| SMAD3 | TGTGAGCTGGATAGACTTGGGAT | CAGATCCTTCCAACTTTTCAAAGTGAAAA | chr15:67483416-67483610 | 2 |
| SMAD3 | CCTCTTTAAAAACTCACTTACGTTTGTCCT | CAACTGACTACATAAACCAATATAGCCGTA | chr15:67483551-67483825 | 1 |
| SMAD3 | GCTTGCTCCAGATTCTGATGCA | CCCTTCTGGTGAATTTGCTTCTGTA | chr15:67483773-67484048 | 2 |
| SMAD3 | CAGCACTTCTGCAAGCTGAAATT | GACTAGTTTTCTCCCTAATTGCTGCT | chr15:67484000-67484259 | 1 |
| SMAD3 | GCAGTTGTATTGTGTAACAAGTACTGCT | CCTTCCCTTCACCTGGAATAAAGC | chr15:67484202-67484476 | 2 |
| SMAD3 | TCAAGGGATTTCCTATGGAAGTCCT | CCCTCCTTCGAGAAGACTCACA | chr15:67484427-67484702 | 1 |
| SMAD3 | AAGGAGCACCTTGACAGACTTG | GCTCTAGCAGAAGCCAAAATGGA | chr15:67484658-67484910 | 2 |
| SMAD3 | CTCTGAGACCCAGGAACCAAATAT | GTAAGACACGACTTCCATAGATCACTG | chr15:67484863-67485087 | 1 |
| SMAD3 | GCACAGCCTGATTCTGGTGATC | GCCTTCAGGTCTGAGCAATGAG | chr15:67485038-67485212 | 2 |
| SMAD3 | GCATTTGGATGGTGTCTTAGAACC | GTTACATACGCCCAAAGCACCTA | chr15:67485166-67485430 | 1 |
| SMAD3 | CATTCCCACCTTAGAGTGCTACA | GTTGACTAGGTGAACGCCACTTA | chr15:67485384-67485615 | 2 |
| SMAD3 | GCTGGTGACTGGATAGCAGTTT | CGTATGTGCTGGAAGCATCTAAGTTA | chr15:67485570-67485827 | 1 |
| SMAD3 | CCACCTCGCTTCTTCCCAATTTAG | CACAGACAACATAAGGACAATGGAGA | chr15:67485777-67486027 | 2 |
| SMAD3 | GACTCTTAGGAAGAGCACACATGAG | TGCCATGCACACACTACTTCAA | chr15:67485956-67486230 | 1 |
| SMAD3 | AATGCTTTTACTCAAGACTACAGAAAGGT | TAAGAAACAGGGTCCAGGGCTAA | chr15:67486179-67486452 | 2 |
| SMAD3 | ATACCAGCAGCAAGAGAGTGCA | CCTCTTGCGTACGTTTTTCTGC | chr15:67486402-67486677 | 1 |
| SMAD3 | GCTTCTCAAGTGGGTAGGGAAA | CATTGCAGAATCACTGAGAATTTCTCA | chr15:67486633-67486833 | 2 |
| SMAD3 | AGAAACTTGCCTCATGTAAACTGGAT | GTTTCTGGATGCTGAGCCAAAAAG | chr15:67486780-67487055 | 1 |
| SMAD3 | GAATGTTGGTGGAGGGTGTAGT | ACACACATCAGAGTCCAGAACAG | chr15:67487007-67487267 | 2 |
| SMAD3 | CAAGAAGCCTTCACTCACCTTCA | TGAGAGGACACAAATCTTTAACAATAGGG | chr15:67487201-67487456 | 1 |
| SMAD3 | CAGAGCACCTGTTTAAGCATTGTAC | GGCTCCCTTTTAAATCTTCTCTCCA | chr15:67487402-67487578 | 2 |
| SMAD2 | GCGGGTATGGAAGACGGAG | GCTTCCCTCGTGCTGATTGG | chr18:45457370-45457555 | 1 |
| SMAD2 | GGACGCACGCAAACACTTC | CCGGGAGTGTTTTCAGTTCC | chr18:45456678-45456872 | 2 |
| SMAD2 | CTCATCTAATCGTCCTGTTTTCTTTAGC | TCAAGGTAGTCTCTACATCATCCTTTCAAT | chr18:45422947-45423221 | 1 |
| SMAD2 | TTCAGTAACTTTCCTTCAGATTCCTTGA | AGCAGTGAAAAGTCTGGTGAAGAA | chr18:45422826-45422999 | 2 |
| SMAD2 | CCCTCCCACAAGATCTCTAAGATTC | CCATGCTTCCATGTTCACATCAATTTTT | chr18:45396799-45396984 | 1 |
| SMAD2 | AGCATATTCGCAGTTTTCAATTGCC | TTTTTAAAGTTCCCTCCTTTCTTTTCCC | chr18:45395677-45395850 | 2 |
| SMAD2 | TTACACTAAAATTTTCCTGGGTCACAAGA | CTGATCTTCACAGTCATCATGAACTCAA | chr18:45395517-45395730 | 1 |
| SMAD2 | CCAAGTTTTAGGAGATTCAGAAGGCA | CTTTGCCATTAACTTGGATTTCTTGAACT | chr18:45394660-45394864 | 2 |
| SMAD2 | ATGCGTCTCAACTTCTCTAAATTTAAGAGT | GGAGTGATTCATTGGTTATCTCTGGT | chr18:45391321-45391594 | 1 |
| SMAD2 | ATTATTTGGCTATTCATTAGGATCCCTTTCTC | AACCAGATAAAGCTAGTTTTATGATTACTTGA | chr18:45377520-45377779 | 2 |

| | | | | |
|---|---|---|---|---|
| SMAD2 | TCTTGTCATTTCTACCGTGGCATT | TGAATTGTTCACGCATTTCTTCCTTTTATT | chr18:45374855-45375113 | 1 |
| SMAD2 | GAAAGCTGGTTTTACTGCACACA | CTTAGGTTTACTCTCCAATGTTAACCGA | chr18:45374739-45374907 | 2 |
| SMAD2 | TAGCAACAAGAAAAACTAAGCAAGTTGAC | CTTCCAAAGTCACACTGAAATAGTAAATGT | chr18:45371971-45372234 | 1 |
| SMAD2 | CCCACCCTTTCACAAAACTCATT | CATGCTCATATTCTAAAACTTGTAACCCTT | chr18:45371723-45371947 | 2 |
| SMAD2 | GCTCAGTAATTTGCAACAGTTTTGAATT | TCAGCTAACTAGAATGTGCACCATAAG | chr18:45371535-45371773 | 1 |
| SMAD2 | ACCACACACAATGCTATGACAGA | GCCTGTGGACTTGAATTTCATAATCATTTT | chr18:45368128-45368370 | 2 |
| SMAD2 | ACTCAGTTTTATGCCCAATAAGACATCATT | TCCCATGAAAAGACTTAATGTAACAACTCT | chr18:45367930-45368181 | 1 |
| SMAD2 | TAACAGAGAAGTGGGAATAACAGATTAGGT | CTCTCCCATTAAAGAGGAAGAGAAGATTTT | chr18:45367738-45367998 | 2 |
| SMAD2 | ACCACAAAAAGAATGACTGTTTAAGCC | TCTAGGCAAACCTTTATAAAGTTGCAGT | chr18:45367543-45367796 | 1 |
| SMAD2 | ACAAAGGGACTTTCTCCATATGAATTCTT | GAAATCCTTGACCTTAATGTGTTCAGTG | chr18:45367326-45367599 | 2 |
| SMAD2 | CCTGAAAATAATTATACTGCCACCTATGCA | GCACCAGTTAATTCTGGGATACAGT | chr18:45367216-45367380 | 1 |
| SMAD2 | CATCACTTTTCTAGGTATTTTGCAGAAACA | GTCTGTTGATTTATCCTTCCACCTCATAAA | chr18:45367011-45367279 | 2 |
| SMAD2 | CATGAAATATTGTCAATCACAAAACCAGGA | GCATTATGAAATGGGCCTTTTGTAAATGTA | chr18:45366799-45367073 | 1 |
| SMAD2 | GAACAAGTACCAAAGACAGCTTCAAAAAT | GCAGTCTGAGCTGTTCTTAACTGA | chr18:45366605-45366853 | 2 |
| SMAD2 | TGTTCACCTTTACCCAAAGACTAAGAA | ACTCCTGGTCGTCTTTTCATAAATTTTCAT | chr18:45366390-45366664 | 1 |
| SMAD2 | CTCCTCACTTGGCTTGCTTCTTTA | TTCACCTGTTTGTACCAGGAACTT | chr18:45366190-45366441 | 2 |
| SMAD2 | CAAAGGTTGAGGAAGGAGATATGTTAGT | ATGCATGAGCTAACACCTGACAT | chr18:45365962-45366237 | 1 |
| SMAD2 | CTGAGTATCTCCTACAGGCCTGAT | TGTGATAGTCTGCTTCATGTGATTCTC | chr18:45365764-45366017 | 2 |
| SMAD2 | GTCCCAATTTCATACTGTACAGAAAAATCG | GCAGAGTAAATGTCCTAGGTAATACCACT | chr18:45365543-45365817 | 1 |
| SMAD2 | AAACATCATCTCGTTAAAAAGCATCAAACA | AGCGCATCTCTACAGTTCTGATG | chr18:45365432-45365596 | 2 |
| SMAD2 | TAGAAAGCATGGCCATTCGGTT | TTCCTGGTTGTAATTACATTTGTGTGAATG | chr18:45365218-45365492 | 1 |
| SMAD2 | CACTAAGTCAGTGAGAACTCCAACAG | ACAATACTGTTAATTTTCTAGGAATGATGACCT | chr18:45365118-45365276 | 2 |
| SMAD2 | GGATAAAAAGAGCACAATCAGCGGTA | AGTAGGTTCCACAGAGACACCTT | chr18:45364950-45365187 | 1 |
| SMAD2 | CAAAGTGACAGGTCCTGAGGAAA | TGGATTTAGGCCTACCTTCTGGAA | chr18:45364726-45365000 | 2 |
| SMAD2 | AAAAGGGAAAGCATTTGACAGTGTTT | CTTGGTCTTGTAGAGGTTGTGTTTTT | chr18:45364571-45364775 | 1 |
| SMAD2 | AAGAGGTAGAGAAACCAACTGGGTATAT | TTGAGTTTTGCTTGATTTGTCTGTCTTTT | chr18:45364417-45364626 | 2 |
| SMAD2 | ATGTACAGTGAAGATTGTCTTGTGTTACTT | CAGTCTTAGTCTCTTGCTAGTTGAAATCAA | chr18:45364261-45364487 | 1 |
| SMAD2 | TAAGTATCAATGCCTTCGATCAGAGTAAGA | GTGGCGTAGGTTTATGTTGGGATATA | chr18:45364099-45364317 | 2 |
| SMAD2 | AAAAACAAGGCTAACAAGAAAGAGGATA | GTCTGTGGACATGAACAATGAATACTTTTT | chr18:45363952-45364159 | 1 |
| SMAD2 | CCCAAGATGGAAATCTGGTATCG | TCTGCATATATCAGAATGTTTCTCTCCTTTG | chr18:45363376-45363637 | 2 |
| SMAD2 | GCTCATATTTCCTGGTTGTAAGGCT | AGTTGCAGCCAATGTTTTCCCTA | chr18:45363158-45363422 | 1 |
| SMAD2 | CCCAACTGTTAAGTTTGTCTATGAGTGTAT | CTTGGAGTGGAATTCCTGTGTTTTG | chr18:45362951-45363208 | 2 |
| SMAD2 | TTAAATGAACTGCCAAATGGCAGATTAAG | ACTGCCTCAGTCTTTTTAAGAAGGAAATAA | chr18:45362802-45363042 | 1 |
| SMAD2 | GAGGGAGCAATCTAGTATTATCAACAACA | TGGACAGACCATTGGAGTACATTAATTTT | chr18:45362631-45362860 | 2 |
| SMAD2 | CAAACCAACTGGCACTGAAGTT | GGGATGTACTAACTTAGGGTTAACTTTGT | chr18:45362420-45362689 | 1 |
| SMAD2 | GAGCATATTTGGAGAAACTAAGAGCTGTTA | AGGTGATCTCGCTGGAGTATATTAACTT | chr18:45362213-45362470 | 2 |
| SMAD2 | ATGGCAAGTTACCTAGTGATTAACATTGT | AGAAAAGTATTCTTTTACCCTCCAAACAGT | chr18:45362009-45362273 | 1 |
| SMAD2 | TTTTTCAGCTCGAACAATCCAAGATT | CTTTCTCTGTGGCTTTAAAGTTTTTGACTA | chr18:45361811-45362072 | 2 |
| SMAD2 | GGTTCTATATTTTGTCAAACAGTTGGGTTT | CATTTGTTTGCCACTCTTACTATTCCC | chr18:45361610-45361864 | 1 |
| SMAD2 | GCCTACAATGCTGTGATCTAGTTCA | CAAGTAGGTAACATGCATGAGTTTTGAATT | chr18:45361402-45361677 | 2 |
| SMAD2 | AAAATATTCTATGCCAACTGTTTTCCACAG | AAGATGAGAACAGTTCTGCAGTTGT | chr18:45361214-45361452 | 1 |
| SMAD2 | AGTAATCAAGCAAATTAGTGAGCAGTGTAT | GCAGCCTTTGTTTTAATATATAATGCAGGA | chr18:45361046-45361274 | 2 |
| SMAD2 | AGAACACTACCCACTCGTTCCTTAATA | CTGAAAACAGATTTCAAGAGTGAGCTTTT | chr18:45360858-45361105 | 1 |
| SMAD2 | TGATGTGCTACTTATCAGAAAAATCCACAT | CCTTGAAAGTTCTTACCTTTTCTCTAGTGA | chr18:45360732-45360915 | 2 |
| SMAD2 | GCTGAGTTTAGAAGCAACTATGACAACA | CCCTGGATACAAAACTTTTAGCAGAGTTT | chr18:45360524-45360798 | 1 |
| SMAD2 | CACATAGGTTCAGAAGAACTTAGCTATCT | GTGTTCTTGTGATTTATCCTTACTTCCCTA | chr18:45360307-45360582 | 2 |
| SMAD2 | CATTCTGGCTTCTCGAGCAGAA | TATAATGGAACAGAGAAGAGGCCAACT | chr18:45360129-45360363 | 1 |
| SMAD2 | GAACAGGGTCTGCATCCATCATA | CGTCTACTGCATTTCCCAGTCT | chr18:45359903-45360174 | 2 |
| SMAD2 | TGTAGGTGGGAGAAAGACATCAGTAT | CTCTGATGGCATTAACTTTGTAATGCAA | chr18:45359695-45359954 | 1 |
| SMAD2 | ACAGGTAAGAGCAAACAAGGTAAAAAGT | ACAGCTGGTTTTCAAGTTAAAATTGTTGA | chr18:45359545-45359750 | 2 |
| SMAD2 | AAAATTAGTCCCATAGTAACATGGTAAACAACT | GTGTGCATCATCTGTATGTGCTTTC | chr18:45359449-45359598 | 1 |
| TGFBR1 | TTTTTCTAAGAATCTTTCTCTTTTTCCAGC | TTCAGCTATACACATGCTGTTGTGTAT | chr9:101891107-101891261 | 1 |
| TGFBR1 | GTCACAGAGACCACAGACAAAGTT | CCAGTTCTAAAATCACAGAGTATGAAGAGT | chr9:101891210-101891453 | 2 |
| TGFBR1 | GGAATTGTGTAGGATTGGGAAAATGG | CCATCAACATGAGTGAGATGCAGA | chr9:101894701-101894880 | 1 |
| TGFBR1 | CTGGACCAGTGTGCTTCG | AGATGTCTTAGGAAAAAGGAGAAACAATTATGT | chr9:101894838-101895057 | 2 |
| TGFBR1 | TCTGGGTCACTCATTAGTGCCT | TCCAAACTTCTCCAAATCGACCTT | chr9:101900054-101900227 | 1 |
| TGFBR1 | CTATTGTGTTACAAGAAAGCATTGGCA | GATGGTTAGTCACCATATCTGTAAAGACTT | chr9:101900176-101900449 | 2 |
| TGFBR1 | GCCCAACCGAAATGTTAATTCTGT | GCCTCCACCTTCTATTTTCATAGACATTAT | chr9:101904786-101905061 | 1 |
| TGFBR1 | TGTAGTGATTCACTTGAGTTTAATAATGCC | AGCAAGTTCCATTCTTCTTTACCAAGATAT | chr9:101906850-101907082 | 2 |
| TGFBR1 | CCATTGCTCATAGAGATTTGAAATCAAAGA | GTTTAAGCTGAGTTTCAGCAATGATATGT | chr9:101907022-101907266 | 1 |

| | | | | |
|---|---|---|---|---|
| TGFBR1 | GAAATGTCTGAAAGGAGGTTCATCCA | AGAAGACAATTCTTGAACAACTTCTGCT | chr9:101908687-101908955 | 2 |
| TGFBR1 | AGGTGATCTTTTAATGCCTTGGCAT | AAAGTAGCAAACTTTTGCTTACTAAGCAG | chr9:101909863-101910138 | 1 |
| TGFBR1 | AATTCTTATCCAGACCAATGGAAAATGG | AAAGCTGTAGAATTACATTTTGATGCCTTC | chr9:101911396-101911599 | 2 |
| TGFBR1 | AAAACATTATCGCAACTCAGTCAACAG | CATTTTCTGTCCTGGGAAAGAAGC | chr9:101911542-101911816 | 1 |
| TGFBR1 | GGGTCCTTTCTGTGCACTATGAAC | GGAAACTCTAGTGGTTCAGAATCCTC | chr9:101911768-101912028 | 2 |
| TGFBR1 | AGTGATTTACTCCTGGTTAGTACATTCTCA | GCACTTACTGGTATAGTACAATCCCATTT | chr9:101911972-101912237 | 1 |
| TGFBR1 | TCACTTATTCAGAACATTACATGCCTTCA | CTTCATTTCTTTAGTGGCTTAAGCACATTT | chr9:101912179-101912451 | 2 |
| TGFBR1 | CTATTCTGAAAATGCCTTTCTCCTACCA | GGCTTACAGAATACCACATAACTTTTCAC | chr9:101912393-101912666 | 1 |
| TGFBR1 | GGGAAAGTCTGTCTAGCTGCTT | ATGTGACTTTCTTGCACATCTGTTATCTAT | chr9:101912615-101912819 | 2 |
| TGFBR1 | CTAGGAAGGCGAAGGTAGTTAATAATTTGA | GGTGCACTGAATGCATTACATAAATTACAT | chr9:101912759-101913000 | 1 |
| TGFBR1 | TGCGTACATTGCAACTGCTTAC | CAGTGATAAAAGGACTTCGAAAACTGT | chr9:101912948-101913207 | 2 |
| TGFBR1 | CTATGAAGTCTCTGCAGGGCTTTT | ACATTATCAGAATGCAACTCATTCAGTGTA | chr9:101913156-101913371 | 1 |
| TGFBR1 | GTCAGGAAGAGAAAGTGGCCATT | TCTCAAACTCATTCATTTGCTTCAATGAAA | chr9:101913318-101913588 | 2 |
| TGFBR1 | CATCATGGGAAAAATGCTTAGAGGTTACTA | GATTTCATAAAGCAGATCTGGTAGGCTT | chr9:101913487-101913743 | 1 |
| TGFBR1 | TTGTTGTTGTTTTTGGGCCATTTCT | AAAAATATCTTCAGCAAAAGTGGATAACCT | chr9:101913690-101913881 | 2 |
| TGFBR1 | AAAGTTGTAATACATGATTTCTCACTTTCATGT | GTTTCTTTCATTTTCAATATCAGCCAATTGTG | chr9:101913817-101914077 | 1 |
| TGFBR1 | TTTTTGGCTAGGGATGGTTTGATAAAC | TTTCTCCTGGCTACTGACAAAATGAAT | chr9:101914018-101914271 | 2 |
| TGFBR1 | TCTTCTGGTAAATTGTGCGTCCAT | CCTCTAATGACTGAAGGAAATGGAAGTG | chr9:101914220-101914492 | 1 |
| TGFBR1 | GTAAAACGGCTGATGGAAGTTGTG | ACCTTACCCTGTCTTATAGGCGA | chr9:101914435-101914662 | 2 |
| TGFBR1 | CCAGAAACAGTGGCCAGTTGTA | GCCAAGTGACCACTTACATTTTGTAATAAA | chr9:101914617-101914796 | 1 |
| TGFBR1 | CCATCAAGAATCCCAGATTTCAGGT | ACGATTCTTCCAGTGCAAGAATAAAAATAA | chr9:101914741-101914956 | 2 |
| TGFBR1 | ACTATCCCATTAGACACATCATCAAAAGC | CTAGAAGTCCAGCACTCTTGAGAG | chr9:101914897-101915163 | 1 |
| TGFBR1 | TCTGTTTTGGATTACTGGAATACCCATG | CGTTGGCACAACGTGAAAAAGG | chr9:101915107-101915378 | 2 |
| TGFBR1 | GTGAACTGAATATCATGAACCATGTTTTGA | GAGTAACTGAGCAACATAATACCCATACTT | chr9:101915322-101915574 | 1 |
| TGFBR1 | CATTCTCCAGGTTTGCAGTTTTATTTCTAT | TCTCAGTATCATTCGACTTCAATGGAAATT | chr9:101915513-101915747 | 2 |
| TGFBR1 | GCTTTATCAGTGTAATCTCTGCCTTTTAAG | GCACTGCTTCCATTCTTCTACTGTAG | chr9:101915663-101915909 | 1 |
| TGFBR1 | GAAGAGTGATGCTTATGTTAAGTCCTAACA | CTAGTTGCCATCTACAGCAACTACAT | chr9:101915853-101916011 | 2 |
| TGFBR1 | ATACGTTGGAATGAGTCATGCCAT | CTGACCCAAAGGCAACAGAGATCAC | chr9:101915961-101916215 | 1 |
| TGFBR1 | GGCATGATGGACCTGTCTACAG | AATCCAGCCTTCAAATTGAAGTAACTATCT | chr9:101916168-101916374 | 2 |
| TGFBR1 | GCTTTGTGAATAGGATTGCTCTCACATTAA | ACGTCCATAGGCAACAAATTGTCTT | chr9:101916314-101916503 | 1 |
| TGFBR2 | GGGCTGGTCTAGGAAACATGA | GCGCGAGTGACTCACTCA | chr3:30647936-30648100 | 1 |
| TGFBR2 | ACAGGAGCCGGACTCCTGTG | GCTGCTCGTCATAGACCGAG | chr3:30648140-30648365 | 2 |
| TGFBR2 | CGGACTCCTGTGCAGCTTC | CCCGCTGCTCGTCATAGA | chr3:30648148-30648368 | 1 |
| TGFBR2 | TTTCAAAACAGTTTCACTTTCCTGTCATC | AAATTAAGCAGTGAGGGAGCATGA | chr3:30664609-30664834 | 2 |
| TGFBR2 | TCTAATCTGATGTGAAGGAATTATTTTGCC | CACCGTTGTTGTCAGTGACTATCAT | chr3:30686087-30686274 | 1 |
| TGFBR2 | ATCTTTCTCTCTCCTCAGTTAATAACGAC | CACTGACTGTGTGTACTATGAGAATACATT | chr3:30686220-30686485 | 2 |
| TGFBR2 | CCCTCGCTTCCAATGAATCTCT | GGTCCCACACCCTTAAGAGAAG | chr3:30691730-30691983 | 1 |
| TGFBR2 | ACTTCCTGACAGTACTTACCTACCA | TTCCCAGGTTGAACTCAGCTTC | chr3:30713065-30713281 | 2 |
| TGFBR2 | ATCTTCTACTGCTACCGCGTTAAC | CTGAAGTGTTCTGCTTCAGCTTG | chr3:30713227-30713480 | 1 |
| TGFBR2 | CGCTTTGCTGAGGTCTATAAGGC | GGTGATCAGCCAGTATTGTTTCCC | chr3:30713434-30713650 | 2 |
| TGFBR2 | CTGAGGAGCGGAAGACGGAGTT | CGAGGATATTGGAGCTCTTGAGGT | chr3:30713603-30713834 | 1 |
| TGFBR2 | CATGTGGGAGGCCCAAGAT | GGCCAGGCTCAAGGTAAAGG | chr3:30713774-30713974 | 2 |
| TGFBR2 | TTGTGAAAATAAAAAGGCAGCTGGAATT | GCAACACATGATCTTATTTTGAAGACAAGT | chr3:30715534-30715802 | 1 |
| TGFBR2 | CATCTCACCATGCTCATTTCCTTTG | TCCAGAATTCTCTGCCACCTAAGA | chr3:30729800-30730075 | 2 |
| TGFBR2 | CATGGTGCCCTTTGGATCTCTT | AGAAGAGCTATTTGGTAGTGTTTAGGGA | chr3:30732878-30733098 | 1 |
| TGFBR2 | CGGAGGAGAAGATTCCTGAAGAC | GCTTATCCTATGACAATGTCAAAGGCATA | chr3:30733044-30733302 | 2 |
| TGFBR2 | GGAGTGGGTGACATAGAGCATTC | TCTCCAATAAAACCAATTTCCTGGGAATAT | chr3:30733250-30733514 | 1 |
| TGFBR2 | CCTTGAAAAGAGACAAGGAAAAACATCAA | TGTACTTAGTTCCATGGCCAGAAGA | chr3:30733455-30733725 | 2 |
| TGFBR2 | AATCATTCCCTGCCTAGCACTTC | GGGACATCTGTTCTGATTTTTCTGC | chr3:30733677-30733940 | 1 |
| TGFBR2 | CCCACTTTTTACCTTCATGGGTT | TGCTCCTTTTTAGAAAAGGATGAAGG | chr3:30733892-30734057 | 2 |
| TGFBR2 | CCTGCACTTATGTTACTATTCTCTGCT | GTCATAGTCCTGACAATGATAGTATTGTGT | chr3:30733999-30734233 | 1 |
| TGFBR2 | GGCCTGATGAAGAGGATTTCAACT | GTAAGTGTCTCCACACCTTCACA | chr3:30734179-30734410 | 2 |
| TGFBR2 | ATCTCCAGTCCACGTTCACAAAA | GGCTAACTGAGACCCTTAAAGGAGT | chr3:30734364-30734638 | 1 |
| TGFBR2 | GAAGTTGGCCTTTTATTGGACTAAAGG | GACCTTTGTGTCCCACATTCAAA | chr3:30734583-30734803 | 2 |
| TGFBR2 | CAACCTTTGCAGAAATTACTAGAGAGGA | AATAAAATGAGACCTTCCACCATCCAA | chr3:30734752-30735026 | 1 |
| TGFBR2 | ACTGGTAGTGAGAATATCAGCTCTGT | TGGACCAGTATTCTAGAAACTCACCA | chr3:30734973-30735231 | 2 |
| TGFBR2 | GCTGTTGCCCATTGACCTCTAG | AGTACAGCTGAAGTGTTCCATAAAAGAAT | chr3:30735183-30735457 | 1 |
| TGFBR2 | GACCAAGGAATAACATTCTGTAGTTCCTAA | CTCACCTGAGTATTTTGCTTTATTCAATCT | chr3:30735355-30735599 | 2 |
| TGFBR2 | GTCTCAAGCACTTATTTTTATTCTATGCATTGT | CATAAATTGCAAAATAACCAAATGAGGCAAA | chr3:30735507-30735669 | 1 |

[illegible]

**Supplementary material S3**. **Characteristics of the polymorphisms detected in 275 healthy and PD individuals**. [1]Chromosome positions according to the NCBI database, build GRCh37. New variants, not present in dbSNP v153, are referred to as through their chromosome: bp position and type of variant: (-) deletion; (NN) insertion; and (N) SNP.

| | | | | | | | Function prediction | |
|---|---|---|---|---|---|---|---|---|
| **Gene** | **SNP ID** | **Chromosome position[1]** | **Allele** | **MAF** | **SNV** | **Type** | **SIFT** | **PolyPhen-2** |
| *TGFB2* | rs10482718 | 1: 218518742 | A/G | 0.02182 | SNP | 5'UTR | | |
| *TGFB2* | rs11466365 | 1: 218519028 | C/T | 0.001818 | SNP | 5'UTR | | |
| *TGFB2* | rs148765724 | 1: 218519706 | T/A | 0.001818 | SNP | 5'UTR | | |
| *TGFB2* | 1:218519756 (-) | 1: 218519756 | A/- | 0.01273 | DEL | 5'UTR | | |
| *TGFB2* | 1:218519880 (G1 | 1: 218519880 | G/GT | 0.06727 | INS | 5'UTR | | |
| *TGFB2* | rs113711540 | 1: 218519928 | A/AAAAC | 0.2691 | INS | 5'UTR | | |
| *TGFB2* | 1:218519942 (-) | 1: 218519942 | A/- | 0.01636 | DEL | 5'UTR | | |
| *TGFB2* | rs200186989 | 1: 218519992 | T/- | 0.05455 | DEL | 5'UTR | | |
| *TGFB2* | 1:218520003 (C | 1: 218520003 | C/CT | 0.01273 | INS | 5'UTR | | |
| *TGFB2* | rs780635221 | 1: 218520020 | C/A | 0.04545 | SNP | 5'UTR | | |
| *TGFB2* | rs781099061 | 1: 218520020 | C/CT | 0.02 | INS | 5'UTR | | |
| *TGFB2* | rs748018397 | 1: 218520021 | T/- | 0.001818 | DEL | 5'UTR | | |
| *TGFB2* | rs779785854 | 1: 218520028 | T/C | 0.001818 | SNP | 5'UTR | | |
| *TGFB2* | rs769838909 | 1: 218520033 | T/- | 0.03818 | DEL | 5'UTR | | |
| *TGFB2* | 1:218520038 (-) | 1: 218520038 | A/- | 0.009091 | DEL | 5'UTR | | |
| *TGFB2* | rs200702935 | 1: 218520039 | A/T | 0.18 | SNP | 5'UTR | | |
| *TGFB2* | 1:218520044 (T) | 1: 218520044 | A/T | 0.007273 | SNP | Missense M-> L | M1L | benign |
| *TGFB2* | 1:218520046 (G | 1: 218520046 | G/GT | 0.01273 | INS | Frameshift | | |
| *TGFB2* | rs371241859 | 1: 218520279 | A/G | 0.001818 | SNP | Missense Q-> R | Q79R | benign |
| *TGFB2* | rs10482721 | 1: 218520315 | G/A | 0.005455 | SNP | Missense R->H | R91H | probably damaging |
| *TGFB2* | rs746220497 | 1: 218520414 | C/A | 0.001818 | SNP | Intron | | |
| *TGFB2* | 1:218578501 (-) | 1: 218578501 | T/- | 0.3545 | DEL | Intron | | |
| *TGFB2* | rs10482810 | 1: 218607532 | G/C | 0.003636 | SNP | Missense V->L | V235L | probably damaging |
| *TGFB2* | rs1375664478 ( | 1: 218607609 | C/T | 0.001818 | SNP | Intron | | |
| *TGFB2* | 1:218607759 (T/ | 1: 218607759 | T/TA | 0.07636 | INS | Frameshift | | |
| *TGFB2* | rs976149098 | 1: 218607817 | G/- | 0.03455 | DEL | Intron | | |
| *TGFB2* | 1:218607828 (-) | 1: 218607828 | G/- | 0.007273 | DEL | Intron | | |
| *TGFB2* | rs774207422 | 1: 218607831 | T/C | 0.001818 | SNP | Intron | | |
| *TGFB2* | rs10482812 | 1: 218607861 | T/C | 0.007273 | SNP | Intron | | |
| *TGFB2* | rs1286186794 | 1: 218609371 | A/- | 0.007273 | DEL | Frameshift | | |
| *TGFB2* | rs767012850 | 1: 218610662 | C/CT | 0.2564 | INS | Intron | | |
| *TGFB2* | rs11285412 | 1: 218610663 | T/- | 0.3273 | DEL | Intron | | |
| *TGFB2* | rs147678881 | 1: 218610736 | G/A | 0.001818 | SNP | Synonymous R->R | | |
| *TGFB2* | 1:218614525 (C) | 1: 218614525 | T/C | 0.001818 | SNP | Intron | | |
| *TGFB2* | rs11466412 | 1: 218614743 | G/A | 0.003636 | SNP | 3'UTR | | |
| *TGFB2* | 1:218614780 (A) | 1: 218614780 | G/A | 0.001818 | SNP | 3'UTR | | |
| *TGFB2* | 1:218614852 (G) | 1: 218614852 | A/G | 0.01818 | SNP | 3'UTR | | |
| *TGFB2* | 1:218614899 (-) | 1: 218614899 | A/- | 0.03636 | DEL | 3'UTR | | |
| *TGFB2* | rs900 | 1: 218614905 | A/T | 0.3036 | SNP | 3'UTR | | |
| *TGFB2* | 1:218614962 (T) | 1: 218614962 | A/T | 0.01091 | SNP | 3'UTR | | |
| *TGFB2* | 1:218614963 (AT | 1: 218614963 | A/AT | 0.02545 | INS | 3'UTR | | |
| *TGFB2* | rs1321255058 (/ | 1: 218614964 | T/A | 0.001818 | SNP | 3'UTR | | |
| *TGFB2* | rs1282624256 | 1: 218614964 | T/- | 0.02909 | DEL | 3'UTR | | |
| *TGFB2* | 1:218614983 (TA | 1: 218614983 | T/TA | 0.12 | INS | 3'UTR | | |
| *TGFB2* | 1:218615025 (G/ | 1: 218615025 | CAACAG/GACAAG | 0.05455 | MNP | 3'UTR | | |
| *TGFB2* | 1:218615203 (G1 | 1: 218615203 | G/GT | 0.01455 | INS | 3'UTR | | |
| *TGFB2* | 1:218615438 (-) | 1: 218615438 | A/- | 0.007273 | DEL | 3'UTR | | |
| *TGFB2* | rs991967 | 1: 218615451 | A/C | 0.3164 | SNP | 3'UTR | | |
| *TGFB2* | rs545208462 | 1: 218615493 | T/- | 0.4436 | DEL | 3'UTR | | |

| | | | | | | | | |
|---|---|---|---|---|---|---|---|---|
| *TGFB2* | rs56271792 | 1: 218615832 | G/A | 0.02545 | SNP | 3'UTR | | |
| *TGFB2* | rs752019003 | 1: 218615835 | T/C | 0.001818 | SNP | 3'UTR | | |
| *TGFB2* | rs532376886 | 1: 218615927 | A/AT | 0.01636 | INS | 3'UTR | | |
| *TGFB2* | rs532935567 | 1: 218615928 | T/- | 0.3836 | DEL | 3'UTR | | |
| *TGFB2* | 1:218615929 (A) | 1: 218615929 | T/A | 0.001818 | SNP | 3'UTR | | |
| *TGFB2* | rs143593418 | 1: 218615977 | T/C | 0.001818 | SNP | 3'UTR | | |
| *TGFB2* | rs1418556 | 1: 218616014 | C/T | 0.2818 | SNP | 3'UTR | | |
| *TGFB2* | rs201094302 | 1: 218616014 | C/- | 0.04364 | DEL | 3'UTR | | |
| *TGFB2* | 1:218616143 (G) | 1: 218616143 | A/G | 0.001818 | SNP | 3'UTR | | |
| *TGFB2* | rs17500758 | 1: 218616206 | T/C | 0.04364 | SNP | 3'UTR | | |
| *TGFB2* | rs3737977 | 1: 218616328 | T/A | 0.07273 | SNP | 3'UTR | | |
| *TGFB2* | rs1245924020 (/ | 1: 218616498 | C/A | 0.001818 | SNP | 3'UTR | | |
| *TGFB2* | 1:218616750 (-) | 1: 218616750 | T/- | 0.01636 | DEL | 3'UTR | | |
| *TGFB2* | rs17047869 | 1: 218616892 | T/C | 0.003636 | SNP | 3'UTR | | |
| *TGFB2* | rs1442089565 | 1: 218616892 | T/- | 0.009091 | DEL | 3'UTR | | |
| *TGFB2* | rs753394620 | 1: 218616910 | T/C | 0.001818 | SNP | 3'UTR | | |
| *TGFB2* | rs1313656124 | 1: 218617051 | G/GT | 0.2491 | INS | 3'UTR | | |
| *TGFB2* | rs3054943 | 1: 218617052 | T/- | 0.2664 | DEL | 3'UTR | | |
| *TGFB2* | rs1046017 | 1: 218617135 | C/G | 0.3139 | SNP | 3'UTR | | |
| *TGFB2* | rs77808075 | 1: 218617268 | A/G | 0.001818 | SNP | 3'UTR | | |
| *TGFB2* | rs746790035 | 1: 218617332 | G/A | 0.001818 | SNP | 3'UTR | | |
| *TGFB2* | rs6704255 | 1: 218617543 | G/A | 0.3109 | SNP | 3'UTR | | |
| *TGFB2* | 1:218617843 (-) | 1: 218617843 | A/- | 0.07636 | DEL | 3'UTR | | |
| *TGFB2* | rs6683598 | 1: 218617900 | C/T | 0.3127 | SNP | 3'UTR | | |
| *TGFBR2* | rs1057139879 | 3: 30647979 | G/A | 0.009091 | SNP | upstream | | |
| *TGFBR2* | rs749340193 | 3: 30648069 | C/T | 0.001818 | SNP | 5'UTR | | |
| *TGFBR2* | rs2306856 | 3: 30648248 | C/G | 0.001818 | SNP | 5'UTR | | |
| *TGFBR2* | rs1155705 | 3: 30686414 | A/G | 0.3309 | SNP | intron | | |
| *TGFBR2* | rs754721647 | 3: 30686423 | A/- | 0.001818 | DEL | intron | | |
| *TGFBR2* | rs756007977 | 3: 30691871 | G/GA | 0.08545 | INS | Frameshift | | |
| *TGFBR2* | rs752580104 | 3: 30691872 | A/- | 0.3855 | DEL | Frameshift | | |
| *TGFBR2* | rs11466512 | 3: 30713126 | T/A | 0.3473 | SNP | intron | | |
| *TGFBR2* | rs2229102 | 3: 30713674 | A/G | 0.005455 | SNP | Synonymous L->L | | |
| *TGFBR2* | rs755843367 | 3: 30713723 | C/T | 0.001818 | SNP | Synonymous L->L | | |
| *TGFBR2* | rs113194608 | 3: 30713737 | C/T | 0.001818 | SNP | Synonymous L->L | | |
| *TGFBR2* | rs35719192 | 3: 30713794 | G/A | 0.005455 | SNP | Missense M->I | M373I | benign |
| *TGFBR2* | rs35766612 | 3: 30713834 | G/A | 0.007273 | SNP | Missense V->M | V387M | probably damaging |
| *TGFBR2* | rs2228048 | 3: 30713842 | C/T | 0.005455 | SNP | Synonymous N->N | | |
| *TGFBR2* | rs2228047 | 3: 30715608 | A/G | 0.003636 | SNP | Synonymous A->A | | |
| *TGFBR2* | rs149112005 | 3: 30733196 | G/A | 0.001818 | SNP | 3'UTR | | |
| *TGFBR2* | rs143024112 | 3: 30733240 | A/T | 0.001818 | SNP | 3'UTR | | |
| *TGFBR2* | rs199931498 | 3: 30733399 | T/- | 0.003636 | DEL | 3'UTR | | |
| *TGFBR2* | rs35934901 | 3: 30733402 | TA/- | 0.5 | DEL | 3'UTR | | |
| *TGFBR2* | rs304840 | 3: 30733421 | C/A | 0.007273 | SNP | 3'UTR | | |
| *TGFBR2* | 3:30733815 (-) | 3: 30733815 | G/- | 0.05636 | DEL | 3'UTR | | |
| *TGFBR2* | rs11466531 | 3: 30733838 | C/G | 0.04182 | SNP | 3'UTR | | |
| *TGFBR2* | rs17026332 | 3: 30733926 | C/A | 0.02545 | SNP | 3'UTR | | |
| *TGFBR2* | 3:30734109 (C) | 3: 30734109 | A/C | 0.001818 | SNP | 3'UTR | | |
| *TGFBR2* | 3:30734410 (T) | 3: 30734410 | C/T | 0.001818 | SNP | 3'UTR | | |
| *TGFBR2* | rs139938413 | 3: 30734598 | A/G | 0.001818 | SNP | 3'UTR | | |
| *TGFBR2* | rs190586104 | 3: 30734653 | T/C | 0.001818 | SNP | 3'UTR | | |
| *TGFBR2* | 3:30734697 (G) | 3: 30734697 | T/G | 0.001818 | SNP | 3'UTR | | |
| *TGFBR2* | rs6550008 | 3: 30734900 | A/G | 0.005455 | SNP | 3'UTR | | |
| *TGFBR2* | rs146296952 | 3: 30734906 | C/T | 0.001818 | SNP | 3'UTR | | |
| *TGFBR2* | rs1803446 | 3: 30734924 | A/C | 0.02909 | SNP | 3'UTR | | |

| | | | | | | | | |
|---|---|---|---|---|---|---|---|---|
| *TGFBR2* | rs188109487 | 3: 30735078 | T/A | 0.001818 | SNP | 3'UTR | | |
| *TGFBR2* | rs1054883228 | 3: 30735112 | T/G | 0.001818 | SNP | 3'UTR | | |
| *TGFBR2* | rs11466536 | 3: 30735156 | C/T | 0.06 | SNP | 3'UTR | | |
| *TGFBR2* | rs11466537 | 3: 30735176 | T/A | 0.005455 | SNP | 3'UTR | | |
| *TGFBR2* | 3:30735319 (GT | 3: 30735319 | G/GT | 0.05818 | INS | 3'UTR | | |
| *TGFBR2* | 3:30735324 (TT | 3: 30735324 | T/TTG | 0.009091 | INS | 3'UTR | | |
| *TGFBR2* | rs947525889 | 3: 30735324 | G/T | 0.009091 | SNP | 3'UTR | | |
| *TGFBR2* | rs542768261 | 3: 30735330 | G/GT | 0.2109 | INS | 3'UTR | | |
| *TGFBR2* | rs1282623826 | 3: 30735331 | T/- | 0.09818 | DEL | 3'UTR | | |
| *TGFBR2* | 3:30735349 (G/ | 3: 30735349 | G/GA | 0.09091 | INS | 3'UTR | | |
| *TGFBR2* | 3:30735350 (-) | 3: 30735350 | T/- | 0.003636 | DEL | 3'UTR | | |
| *TGFBR2* | 3:30735396 (CT | 3: 30735396 | C/CT | 0.02 | INS | 3'UTR | | |
| *TGFBR1* | rs145033378 | 9: 101891246 | C/T | 0.001818 | SNP | Synonymous S->S | | |
| *TGFBR1* | rs1321978114 | 9: 101891397 | A/AT | 0.03818 | INS | Intron | | |
| *TGFBR1* | rs56014374 | 9: 101894904 | G/A | 0.02 | SNP | Missense V->I | V153I | benign |
| *TGFBR1* | rs7861780 | 9: 101907165 | A/C | 0.007273 | SNP | Synonymous T->T | | |
| *TGFBR1* | rs746666082 | 9: 101908774 | G/T | 0.001818 | SNP | Missense A->S | A380S | probably damaging |
| *TGFBR1* | rs863223825 | 9: 101908802 | A/G | 0.001818 | SNP | Missense N->S | N393S | probably damaging |
| *TGFBR1* | rs334354 | 9: 101908915 | G/A | 0.1873 | SNP | Intron | | |
| *TGFBR1* | 9:101908916 (-) | 9: 101908916 | T/- | 0.005455 | DEL | Intron | | |
| *TGFBR1* | rs766851280 | 9: 101909916 | T/- | 0.001818 | DEL | Intron | | |
| *TGFBR1* | rs752215820 | 9: 101911436 | A/AT | 0.3727 | INS | Intron | | |
| *TGFBR1* | rs778258700 | 9: 101911437 | T/- | 0.1673 | DEL | Intron | | |
| *TGFBR1* | 9:101911613 (CT) | 9: 101911613 | C/CT | 0.03636 | INS | 3'UTR | | |
| *TGFBR1* | rs1473423734 | 9: 101911630 | C/T | 0.001818 | SNP | 3'UTR | | |
| *TGFBR1* | rs868 | 9: 101911656 | A/G | 0.1836 | SNP | 3'UTR | | |
| *TGFBR1* | 9:101911730 (T) | 9: 101911730 | A/T | 0.001818 | SNP | 3'UTR | | |
| *TGFBR1* | rs145692006 | 9: 101911947 | A/G | 0.003636 | SNP | 3'UTR | | |
| *TGFBR1* | rs201269197 | 9: 101912089 | T/C | 0.001818 | SNP | 3'UTR | | |
| *TGFBR1* | 9:101912090 (TA | 9: 101912090 | T/TA | 0.003636 | INS | 3'UTR | | |
| *TGFBR1* | rs866132758 | 9: 101912151 | A/G | 0.001818 | SNP | 3'UTR | | |
| *TGFBR1* | rs201687617 | 9: 101912161 | G/A | 0.003636 | SNP | 3'UTR | | |
| *TGFBR1* | rs1315984960 | 9: 101912440 | A/G | 0.001818 | SNP | 3'UTR | | |
| *TGFBR1* | rs334348 | 9: 101912471 | A/G | 0.2618 | SNP | 3'UTR | | |
| *TGFBR1* | rs923536988 | 9: 101912667 | A/AT | 0.2899 | INS | 3'UTR | | |
| *TGFBR1* | rs200689459 | 9: 101912827 | G/A | 0.001818 | SNP | 3'UTR | | |
| *TGFBR1* | rs11568811 | 9: 101913015 | G/C | 0.009091 | SNP | 3'UTR | | |
| *TGFBR1* | rs760928585 | 9: 101913042 | T/C | 0.001818 | SNP | 3'UTR | | |
| *TGFBR1* | rs780605360 | 9: 101913273 | TA/- | 0.001818 | DEL | 3'UTR | | |
| *TGFBR1* | rs181403208 | 9: 101913284 | C/T | 0.007273 | SNP | 3'UTR | | |
| *TGFBR1* | rs201191009 | 9: 101913359 | A/C | 0.001818 | SNP | 3'UTR | | |
| *TGFBR1* | 9:101913393 (TA | 9: 101913393 | T/TA | 0.02727 | INS | 3'UTR | | |
| *TGFBR1* | 9:101913536 (GT | 9: 101913536 | G/GT | 0.07273 | INS | 3'UTR | | |
| *TGFBR1* | 9:101913654 (CT | 9: 101913654 | C/CT | 0.02545 | INS | 3'UTR | | |
| *TGFBR1* | rs879166510 | 9: 101913668 | G/GT | 0.05273 | INS | 3'UTR | | |
| *TGFBR1* | rs75796733 | 9: 101913669 | T/- | 0.01273 | DEL | 3'UTR | | |
| *TGFBR1* | rs201152885 | 9: 101913678 | T/G | 0.01636 | SNP | 3'UTR | | |
| *TGFBR1* | 9:101913678-10 | 9: 101913678-101913680 | TTTG/TGTT | 0.007273 | MNP | 3'UTR | | |
| *TGFBR1* | rs199545273 | 9: 101913679 | G/T | 0.2127 | SNP | 3'UTR | | |
| *TGFBR1* | rs202187266 | 9: 101913680 | T/G | 0.007273 | SNP | 3'UTR | | |
| *TGFBR1* | rs532530728 | 9: 101913687 | T/G | 0.009091 | SNP | 3'UTR | | |
| *TGFBR1* | 9:101913688 (G) | 9: 101913688 | T/G | 0.009091 | SNP | 3'UTR | | |
| *TGFBR1* | 9:101913689 (G) | 9: 101913689 | T/G | 0.02 | SNP | 3'UTR | | |
| *TGFBR1* | rs886063239 | 9: 101913689 | T/TG | 0.001818 | INS | 3'UTR | | |
| *TGFBR1* | rs7871490 | 9: 101913690 | T/G | 0.09636 | SNP | 3'UTR | | |

| | | | | | | | | | |
|---|---|---|---|---|---|---|---|---|---|
| *TGFBR1* | rs36064078 | 9: | 101913690 | T/TG | 0.03636 | INS | 3'UTR | | |
| *TGFBR1* | 9:101913692-10 | 9: | 101913692 | TG/GT | 0.005455 | INV | 3'UTR | | |
| *TGFBR1* | rs201054018 | 9: | 101913693 | G/T | 0.02909 | SNP | 3'UTR | | |
| *TGFBR1* | rs1274989626 | 9: | 101913696 | G/T | 0.007273 | SNP | 3'UTR | | |
| *TGFBR1* | 9:101913958 (AT | 9: | 101913958 | A/AT | 0.001818 | INS | 3'UTR | | |
| *TGFBR1* | rs17724567 | 9: | 101914232 | A/C | 0.007273 | SNP | 3'UTR | | |
| *TGFBR1* | rs200274678 (G | 9: | 101914386 | C/G | 0.001818 | SNP | 3'UTR | | |
| *TGFBR1* | rs334349 | 9: | 101914387 | G/A | 0.2636 | SNP | 3'UTR | | |
| *TGFBR1* | 9:101914443 (-) | 9: | 101914443 | G/- | 0.1873 | DEL | 3'UTR | | |
| *TGFBR1* | rs997094252 | 9: | 101914748 | A/G | 0.001818 | SNP | 3'UTR | | |
| *TGFBR1* | rs201114226 | 9: | 101914843 | A/C | 0.001818 | SNP | 3'UTR | | |
| *TGFBR1* | rs117440593 | 9: | 101914848 | T/C | 0.003636 | SNP | 3'UTR | | |
| *TGFBR1* | rs420549 | 9: | 101914873 | G/C | 0.1873 | SNP | 3'UTR | | |
| *TGFBR1* | rs41283640 | 9: | 101915627 | T/A | 0.003636 | SNP | 3'UTR | | |
| *TGFBR1* | 9:101915704 (G/ | 9: | 101915704 | G/GA | 0.01273 | INS | 3'UTR | | |
| *TGFBR1* | 9:101915764 (G/ | 9: | 101915764 | G/GA | 0.02364 | INS | 3'UTR | | |
| *TGFBR1* | 9:101915793 (GT | 9: | 101915793 | G/GT | 0.04396 | INS | 3'UTR | | |
| *TGFBR1* | rs41283642 | 9: | 101915887 | C/T | 0.02364 | SNP | 3'UTR | | |
| *TGFBR1* | rs7850895 | 9: | 101916076 | T/C | 0.09636 | SNP | 3'UTR | | |
| *TGFBR1* | rs41274644 | 9: | 101916080 | G/A | 0.005455 | SNP | 3'UTR | | |
| *TGFBR1* | rs79641064 | 9: | 101916086 | G/A | 0.03091 | SNP | 3'UTR | | |
| *TGFBR1* | rs1590 | 9: | 101916165 | T/G | 0.2673 | SNP | 3'UTR | | |
| *TGFBR1* | rs202059366 | 9: | 101916171 | C/T | 0.001818 | SNP | 3'UTR | | |
| *TGFBR1* | rs200806851 | 9: | 101916210 | G/C | 0.001818 | SNP | 3'UTR | | |
| *TGFBR1* | 9:101916329 (G) | 9: | 101916329 | A/G | 0.001818 | SNP | 3'UTR | | |
| *TGFBR1* | rs561572489 | 9: | 101916380 | A/AT | 0.2382 | INS | 3'UTR | | |
| *TGFBR1* | rs968481994 | 9: | 101916381 | T/- | 0.1 | DEL | 3'UTR | | |
| *TGFBR1* | rs185400896 | 9: | 101916396 | A/T | 0.001818 | SNP | 3'UTR | | |
| *TGFBR1* | rs35118283 | 9: | 101916441 | A/- | 0.06 | DEL | 3'UTR | | |
| *SMAD3* | rs918127087 | 15: | 67357135 | G/A | 0.001818 | SNP | Upstream | | |
| *SMAD3* | rs899894403 | 15: | 67357321 | T/C | 0.009091 | SNP | Upstream | | |
| *SMAD3* | 15:67357411 (-) | 15: | 67357411 | G/- | 0.2545 | DEL | Upstream | | |
| *SMAD3* | 15:67357991 (-) | 15: | 67357991 | C/- | 0.2218 | DEL | Upstream | | |
| *SMAD3* | 15:67357993 (-) | 15: | 67357993 | G/- | 0.01455 | DEL | Upstream | | |
| *SMAD3* | rs1241962534 | 15: | 67358099 | G/A | 0.001818 | SNP | Upstream | | |
| *SMAD3* | 15:67358388 (A | 15: | 67358388 | G/A | 0.001825 | SNP | 5'UTR | | |
| *SMAD3* | rs144374592 | 15: | 67358465 | C/T | 0.00365 | SNP | 5'UTR | | |
| *SMAD3* | rs36221703 | 15: | 67358470 | C/T | 0.001818 | SNP | 5'UTR | | |
| *SMAD3* | rs1061427 | 15: | 67358478 | G/A | 0.2198 | SNP | 5'UTR | | |
| *SMAD3* | rs11636161 | 15: | 67418104 | G/A | 0.3164 | SNP | 5'UTR | | |
| *SMAD3* | rs16950635 | 15: | 67418205 | G/A | 0.06364 | SNP | 5'UTR | | |
| *SMAD3* | rs8028147 | 15: | 67418233 | G/A | 0.2709 | SNP | 5'UTR | | |
| *SMAD3* | rs1172546213 | 15: | 67430390 | C/T | 0.001818 | SNP | Missensen T -> M T9M isoform 2 | | |
| *SMAD3* | rs1065080 | 15: | 67457335 | G/A | 0.1073 | SNP | Synonymous L->L | | |
| *SMAD3* | rs200305159 | 15: | 67457480 | T/TC | 0.04182 | INS | Intron | | |
| *SMAD3* | rs2289261 | 15: | 67457485 | C/G | 0.3636 | SNP | Intron | | |
| *SMAD3* | 15:67457494 (T( | 15: | 67457494 | T/TC | 0.003636 | INS | Intron | | |
| *SMAD3* | rs869167666 | 15: | 67457495 | C/- | 0.007273 | DEL | Intron | | |
| *SMAD3* | rs200689238 | 15: | 67457507 | C/T | 0.003636 | SNP | Intron | | |
| *SMAD3* | rs863223759 | 15: | 67457640 | C/- | 0.07818 | DEL | Frameshift | | |
| *SMAD3* | rs35874463 | 15: | 67457698 | A/G | 0.04727 | SNP | Missensen I -> V | I170V | benign |
| *SMAD3* | 15:67457726 (-) | 15: | 67457726 | G/- | 0.005455 | DEL | intron | | |
| *SMAD3* | rs56336520 | 15: | 67457840 | G/A | 0.02 | SNP | Intron | | |
| *SMAD3* | rs2289259 | 15: | 67457850 | G/A | 0.08182 | SNP | Intron | | |
| *SMAD3* | rs761730786 | 15: | 67458399 | G/A | 0.001818 | SNP | Intron | | |

| | | | | | | | | | |
|---|---|---|---|---|---|---|---|---|---|
| *SMAD3* | rs189793484 | 15: | 67458427 | G/T | 0.001818 | SNP | Intron | | |
| *SMAD3* | rs186178811 | 15: | 67458607 | A/G | 0.001818 | SNP | Intron | | |
| *SMAD3* | rs536010162 | 15: | 67459279 | C/T | 0.007273 | SNP | Intron | | |
| *SMAD3* | rs56070432 | 15: | 67463000 | C/T | 0.02545 | SNP | Intron | | |
| *SMAD3* | rs61258186 | 15: | 67463041 | C/T | 0.003636 | SNP | Intron | | |
| *SMAD3* | rs1278345256 | 15: | 67473676 | G/A | 0.01316 | SNP | Synonymous Q->Q | | |
| *SMAD3* | 1:67473677 (T) | 15: | 67473677 | C/T | 0.01124 | SNP | Intron | | |
| *SMAD3* | 1:67473678 (T) | 15: | 67473678 | C/T | 0.001873 | SNP | Intron | | |
| *SMAD3* | rs1320490523 (/ | 15: | 67473736 | G/A | 0.001818 | SNP | Synonymous G->G | | |
| *SMAD3* | rs984900419 (A) | 15: | 67473742 | C/A | 0.2521 | SNP | Synonymous L->L | | |
| *SMAD3* | rs730880214 | 15: | 67473780 | G/A | 0.008097 | SNP | Missensen R -> Q | R287Q | probably damaging |
| *SMAD3* | rs117185005 | 15: | 67473790 | C/T | 0.01562 | SNP | Synonymous I->I | | |
| *SMAD3* | 15:67473800 (G | 15: | 67473800 | T/G | 0.05921 | SNP | Intron | | |
| *SMAD3* | rs745896273 | 15: | 67473813 | C/T | 0.006522 | SNP | Intron | | |
| *SMAD3* | rs775394029 | 15: | 67473816 | C/T | 0.002165 | SNP | Intron | | |
| *SMAD3* | 15:67473817 (T) | 15: | 67473817 | C/T | 0.01299 | SNP | Intron | | |
| *SMAD3* | 15:67473819 (C' | 15: | 67473819 | C/CT | 0.004329 | INS | Intron | | |
| *SMAD3* | rs761694392 | 15: | 67473820 | C/A | 0.02381 | SNP | Intron | | |
| *SMAD3* | rs200703402 | 15: | 67473821 | C/T | 0.004329 | SNP | Intron | | |
| *SMAD3* | rs536505804 | 15: | 67473822 | A/C | 0.0678 | SNP | Intron | | |
| *SMAD3* | rs776577220 | 15: | 67477061 | G/A | 0.001812 | SNP | intron | | |
| *SMAD3* | rs753875974 | 15: | 67479785 | C/T | 0.003636 | SNP | Synonymous Y->Y | | |
| *SMAD3* | 15:67482775 (T) | 15: | 67482775 | C/T | 0.001818 | SNP | Synonymous P->P | | |
| *SMAD3* | rs368655036 | 15: | 67482925 | G/C | 0.001818 | SNP | 3'UTR | | |
| *SMAD3* | rs534838557 | 15: | 67483029 | C/T | 0.001818 | SNP | 3'UTR | | |
| *SMAD3* | rs576999514 | 15: | 67483089 | G/C | 0.001818 | SNP | 3'UTR | | |
| *SMAD3* | 15:67483167 (-) | 15: | 67483167 | C/- | 0.1145 | DEL | 3'UTR | | |
| *SMAD3* | 15:67483170-67 | 15: | 67483171 | CCTC/CT | 0.02545 | DEL | 3'UTR | | |
| *SMAD3* | rs72661159 | 15: | 67483175 | C/T | 0.001818 | SNP | 3'UTR | | |
| *SMAD3* | 15:67483185 (-) | 15: | 67483185 | T/- | 0.1418 | DEL | 3'UTR | | |
| *SMAD3* | 15:67483233 (A | 15: | 67483233 | A/AC | 0.001818 | INS | 3'UTR | | |
| *SMAD3* | rs8025774 | 15: | 67483276 | C/T | 0.2618 | SNP | 3'UTR | | |
| *SMAD3* | 15:67483294 (-) | 15: | 67483294 | C/- | 0.1255 | DEL | 3'UTR | | |
| *SMAD3* | 15:67483470 (-) | 15: | 67483470 | C/- | 0.04 | DEL | 3'UTR | | |
| *SMAD3* | 15:67483581 (C) | 15: | 67483581 | T/C | 0.001818 | SNP | 3'UTR | | |
| *SMAD3* | rs55970514 | 15: | 67483717 | G/A | 0.01455 | SNP | 3'UTR | | |
| *SMAD3* | rs8031440 | 15: | 67483979 | G/A | 0.2545 | SNP | 3'UTR | | |
| *SMAD3* | rs567052377 | 15: | 67484039 | A/G | 0.001818 | SNP | 3'UTR | | |
| *SMAD3* | rs72661160 | 15: | 67484055 | C/T | 0.005455 | SNP | 3'UTR | | |
| *SMAD3* | 15:67484081 (-) | 15: | 67484081 | G/- | 0.1909 | DEL | 3'UTR | | |
| *SMAD3* | rs8031627 | 15: | 67484119 | G/A | 0.2527 | SNP | 3'UTR | | |
| *SMAD3* | 15:67484290 (C | 15: | 67484290 | G/C | 0.001818 | SNP | 3'UTR | | |
| *SMAD3* | rs2278670 | 15: | 67484297 | C/T | 0.2564 | SNP | 3'UTR | | |
| *SMAD3* | rs144205938 | 15: | 67484754 | C/T | 0.007273 | SNP | 3'UTR | | |
| *SMAD3* | rs549152312 | 15: | 67484902 | T/C | 0.003636 | SNP | 3'UTR | | |
| *SMAD3* | rs11638476 | 15: | 67485101 | G/A | 0.3673 | SNP | 3'UTR | | |
| *SMAD3* | 15:67485102 (A) | 15: | 67485102 | G/A | 0.02364 | SNP | 3'UTR | | |
| *SMAD3* | rs188412401 | 15: | 67485103 | A/G | 0.009091 | SNP | 3'UTR | | |
| *SMAD3* | rs573496525 | 15: | 67485103 | A/- | 0.16 | DEL | 3'UTR | | |
| *SMAD3* | rs12595334 | 15: | 67485156 | C/T | 0.2618 | SNP | 3'UTR | | |
| *SMAD3* | rs755985886 | 15: | 67485277 | C/T | 0.001818 | SNP | 3'UTR | | |
| *SMAD3* | rs72661161 | 15: | 67485421 | C/T | 0.005455 | SNP | 3'UTR | | |
| *SMAD3* | 15:67485565 (C | 15: | 67485565 | A/C | 0.001818 | SNP | 3'UTR | | |
| *SMAD3* | rs3743342 | 15: | 67485667 | C/T | 0.2527 | SNP | 3'UTR | | |
| *SMAD3* | rs145645018 | 15: | 67485911 | C/G | 0.003636 | SNP | 3'UTR | | |

| | | | | | | |
|---|---|---|---|---|---|---|
| *SMAD3* | 15:67485918 (-) | 15: 67485918 | T/- | 0.08 | DEL | 3'UTR |
| *SMAD3* | rs3833027 | 15: 67485934 | AGAG/A | 0.001818 | DEL | 3'UTR |
| *SMAD3* | rs62014610 | 15: 67486000 | C/T | 0.001818 | SNP | 3'UTR |
| *SMAD3* | 15:67486010 (A) | 15: 67486010 | G/A | 0.003636 | SNP | 3'UTR |
| *SMAD3* | rs747191952 | 15: 67486032 | A/AT | 0.1382 | INS | 3'UTR |
| *SMAD3* | rs878890006 | 15: 67486033 | T/- | 0.3873 | DEL | 3'UTR |
| *SMAD3* | rs11556089 | 15: 67486078 | G/A | 0.08727 | SNP | 3'UTR |
| *SMAD3* | rs11556090 | 15: 67486383 | G/A | 0.4836 | SNP | 3'UTR |
| *SMAD3* | rs12900401 | 15: 67486590 | C/T | 0.04364 | SNP | 3'UTR |
| *SMAD3* | rs3743343 | 15: 67486775 | T/C | 0.2655 | SNP | 3'UTR |
| *SMAD3* | rs975954141 | 15: 67486835 | A/AT | 0.1164 | INS | 3'UTR |
| *SMAD3* | 15:67486836 (-) | 15: 67486836 | T/- | 0.1073 | DEL | 3'UTR |
| *SMAD3* | rs1052488 | 15: 67486847 | T/C | 0.2527 | SNP | 3'UTR |
| *SMAD3* | 15:67486933 (A | 15: 67486933 | A/AT | 0.001818 | INS | 3'UTR |
| *SMAD3* | 15:67486934 (-) | 15: 67486934 | T/- | 0.02182 | DEL | 3'UTR |
| *SMAD3* | rs72661162 | 15: 67487275 | T/C | 0.01091 | SNP | 3'UTR |
| *SMAD3* | rs757151117 | 15: 67487324 | C/G | 0.003636 | SNP | 3'UTR |
| *SMAD3* | rs10438355 | 15: 67487549 | C/G | 0.2691 | SNP | downstream |
| *SMAD2* | 18:45456728 (T | 18: 45456728 | C/T | 0.007273 | SNP | Intron |
| *SMAD2* | 18:45456727 (T | 18: 45456727 | C/T | 0.3618 | SNP | Intron |
| *SMAD2* | 18:45456724-4! | 18: 45456724-45456726 | GCT/GGC | 0.3618 | MNP | Intron |
| *SMAD2* | rs552945104 (T) | 18: 45395741 | C/T | 0.001818 | SNP | Synonymous L->L |
| *SMAD2* | 18:45395624 (T | 18: 45395624 | A/T | 0.2436 | SNP | Synonymous V->V |
| *SMAD2* | 18:45391525 (G | 18: 45391525 | T/G | 0.001818 | SNP | Intron |
| *SMAD2* | 18:45391509 (C/ | 18: 45391509 | C/CA | 0.005455 | INS | Intron |
| *SMAD2* | rs149135973 | 18: 45391419 | T/TT | 0.1491 | INS | Intron |
| *SMAD2* | rs1177379668 | 18: 45391390 | T/C | 0.001818 | SNP | Intron |
| *SMAD2* | rs72661147 | 18: 45391356 | T/C | 0.003636 | SNP | Intron |
| *SMAD2* | 18:45377561 (-) | 18: 45377561 | A/- | 0.04364 | DEL | Intron |
| *SMAD2* | 18:45371864 (T/ | 18: 45371864 | T/TA | 0.009091 | INS | Intron |
| *SMAD2* | rs72661148 | 18: 45371695 | A/G | 0.04727 | SNP | Intron |
| *SMAD2* | 18:45371573 (-) | 18: 45371573 | C/- | 0.003636 | DEL | 3'UTR |
| *SMAD2* | rs866244434 | 18: 45368107 | G/A | 0.003636 | SNP | 3'UTR |
| *SMAD2* | rs575364955 | 18: 45367938 | T/G | 0.001818 | SNP | 3'UTR |
| *SMAD2* | rs192433123 | 18: 45367802 | A/C | 0.001818 | SNP | 3'UTR |
| *SMAD2* | 18:45367652 (G | 18: 45367652 | G/GA | 0.01636 | INS | 3'UTR |
| *SMAD2* | 18:45367562 (T | 18: 45367562 | T/TAA | 0.001818 | INS | 3'UTR |
| *SMAD2* | rs1001667150 | 18: 45367537 | A/C | 0.001818 | SNP | 3'UTR |
| *SMAD2* | 18:45367529 (C | 18: 45367529 | A/C | 0.009091 | SNP | 3'UTR |
| *SMAD2* | rs200393944 | 18: 45367528 | C/A | 0.03455 | SNP | 3'UTR |
| *SMAD2* | rs201278324 | 18: 45367521 | AAAAAAAAC/- | 0.06545 | DEL | 3'UTR |
| *SMAD2* | rs1158092324 | 18: 45367517 | A/- | 0.08364 | DEL | 3'UTR |
| *SMAD2* | rs1170854585 | 18: 45367505 | G/T | 0.001818 | SNP | 3'UTR |
| *SMAD2* | rs189397494 | 18: 45367380 | C/T | 0.001818 | SNP | 3'UTR |
| *SMAD2* | 18:45367360 (C | 18: 45367360 | T/C | 0.001818 | SNP | 3'UTR |
| *SMAD2* | rs527529143 | 18: 45367179 | A/C | 0.001818 | SNP | 3'UTR |
| *SMAD2* | rs1241300860 | 18: 45367134 | T/C | 0.001818 | SNP | 3'UTR |
| *SMAD2* | 18:45367103 (C) | 18: 45367103 | T/C | 0.001818 | SNP | 3'UTR |
| *SMAD2* | 18:45366897 (T | 18: 45366897 | C/T | 0.003636 | SNP | 3'UTR |
| *SMAD2* | 18:45366500 (T | 18: 45366500 | C/T | 0.001818 | SNP | 3'UTR |
| *SMAD2* | 18:45366465 (T | 18: 45366465 | T/TA | 0.05636 | INS | 3'UTR |
| *SMAD2* | rs12954768 | 18: 45366422 | T/G | 0.005455 | SNP | 3'UTR |
| *SMAD2* | rs72661150 | 18: 45366210 | C/T | 0.005455 | SNP | 3'UTR |
| *SMAD2* | rs72661151 | 18: 45366140 | C/T | 0.01455 | SNP | 3'UTR |
| *SMAD2* | 18:45366132 (A) | 18: 45366132 | G/A | 0.001818 | SNP | 3'UTR |

| | | | | | | |
|---|---|---|---|---|---|---|
| *SMAD2* | rs72661152 | 18: 45366084 | T/C | 0.005455 | SNP | 3'UTR |
| *SMAD2* | rs560729239 | 18: 45365711 | T/C | 0.003636 | SNP | 3'UTR |
| *SMAD2* | rs149229274 | 18: 45365688 | G/C | 0.005455 | SNP | 3'UTR |
| *SMAD2* | 18:45365683 (C | 18: 45365683 | T/C | 0.001818 | SNP | 3'UTR |
| *SMAD2* | rs5824709 | 18: 45365397 | A/- | 0.3285 | DEL | 3'UTR |
| *SMAD2* | rs1251386714 | 18: 45365396 | C/CA | 0.2318 | INS | 3'UTR |
| *SMAD2* | 18:45365378 (T | 18: 45365378 | T/TG | 0.005455 | INS | 3'UTR |
| *SMAD2* | rs112132589 | 18: 45365336 | G/A | 0.009091 | SNP | 3'UTR |
| *SMAD2* | 18:45365330 (C | 18: 45365330 | C/CA | 0.01091 | INS | 3'UTR |
| *SMAD2* | 18:45365319 (C | 18: 45365319 | C/CA | 0.01091 | INS | 3'UTR |
| *SMAD2* | rs1011483314 | 18: 45365066 | A/T | 0.001818 | SNP | 3'UTR |
| *SMAD2* | rs928231569 | 18: 45365017 | C/T | 0.001818 | SNP | 3'UTR |
| *SMAD2* | rs139145682 | 18: 45365014 | A/G | 0.001818 | SNP | 3'UTR |
| *SMAD2* | rs1478104599 | 18: 45364720 | CTT/- | 0.001818 | DEL | 3'UTR |
| *SMAD2* | rs1450957374 | 18: 45364543 | C/CA | 0.04364 | INS | 3'UTR |
| *SMAD2* | rs375946894 | 18: 45364489 | TGTT/- | 0.007273 | DEL | 3'UTR |
| *SMAD2* | rs17813836 | 18: 45364276 | T/C | 0.009091 | SNP | 3'UTR |
| *SMAD2* | rs142213588 | 18: 45364208 | G/A | 0.01818 | SNP | 3'UTR |
| *SMAD2* | rs183642985 | 18: 45364038 | G/A | 0.007273 | SNP | 3'UTR |
| *SMAD2* | rs117487905 | 18: 45363411 | T/C | 0.001818 | SNP | 3'UTR |
| *SMAD2* | rs8085335 | 18: 45363289 | A/G | 0.05818 | SNP | 3'UTR |
| *SMAD2* | rs1792666 | 18: 45363214 | A/T | 0.4982 | SNP | 3'UTR |
| *SMAD2* | rs146897233 | 18: 45363117 | T/TA | 0.05091 | INS | 3'UTR |
| *SMAD2* | 18:45362911 (G | 18: 45362911 | G/GT | 0.003636 | INS | 3'UTR |
| *SMAD2* | rs16958509 | 18: 45362796 | T/C | 0.005455 | SNP | 3'UTR |
| *SMAD2* | 18:45362790 (-) | 18: 45362790 | A/- | 0.04909 | DEL | 3'UTR |
| *SMAD2* | rs8098413 | 18: 45362721 | C/T | 0.06545 | SNP | 3'UTR |
| *SMAD2* | rs147470332 | 18: 45362509 | T/C | 0.009091 | SNP | 3'UTR |
| *SMAD2* | rs1019023922 | 18: 45362459 | G/A | 0.001818 | SNP | 3'UTR |
| *SMAD2* | 18:45362372 (G | 18: 45362372 | A/G | 0.001818 | SNP | 3'UTR |
| *SMAD2* | rs1050448319 | 18: 45362279 | T/TT | 0.005455 | INS | 3'UTR |
| *SMAD2* | rs547209827 | 18: 45362245 | C/T | 0.001818 | SNP | 3'UTR |
| *SMAD2* | rs1792671 | 18: 45362194 | C/T | 0.4464 | SNP | 3'UTR |
| *SMAD2* | rs180935952 | 18: 45362160 | C/A | 0.001818 | SNP | 3'UTR |
| *SMAD2* | 18:45362136 (C | 18: 45362136 | C/CT | 0.001818 | INS | 3'UTR |
| *SMAD2* | 18:45361809 (-) | 18: 45361809 | T/- | 0.1764 | DEL | 3'UTR |
| *SMAD2* | rs182383364 | 18: 45361643 | A/T | 0.003636 | SNP | 3'UTR |
| *SMAD2* | rs1362163540 | 18: 45361642 | T/A | 0.01091 | SNP | 3'UTR |
| *SMAD2* | 18:45361523 (T | 18: 45361523 | T/TA | 0.02364 | INS | 3'UTR |
| *SMAD2* | rs116123302 | 18: 45361493 | G/C | 0.001818 | SNP | 3'UTR |
| *SMAD2* | 18:45361489 (T | 18: 45361489 | T/TA | 0.02364 | INS | 3'UTR |
| *SMAD2* | 18:45361464 (G | 18: 45361464 | G/GT | 0.01091 | INS | 3'UTR |
| *SMAD2* | rs145901048 | 18: 45361363 | G/A | 0.001818 | SNP | 3'UTR |
| *SMAD2* | rs72912348 | 18: 45361245 | T/C | 0.007273 | SNP | 3'UTR |
| *SMAD2* | rs1981 | 18: 45360991 | G/A | 0.4418 | SNP | 3'UTR |
| *SMAD2* | rs562651074 | 18: 45360860 | G/C | 0.001818 | SNP | 3'UTR |
| *SMAD2* | rs776570464 | 18: 45360801 | A/G | 0.001818 | SNP | 3'UTR |
| *SMAD2* | rs115139672 | 18: 45360184 | G/C | 0.001818 | SNP | 3'UTR |
| *SMAD2* | rs1435018887 | 18: 45359955 | A/- | 0.1127 | DEL | 3'UTR |
| *SMAD2* | rs79514034 | 18: 45359803 | C/T | 0.007273 | SNP | 3'UTR |
| *SMAD2* | rs8671 | 18: 45359664 | A/T | 0.4909 | SNP | 3'UTR |
| *SMAD2* | rs763168619 | 18: 45359618 | C/T | 0.001818 | SNP | 3'UTR |
| *SMAD2* | rs557633152 | 18: 45359568 | A/G | 0.003636 | SNP | 3'UTR |
| [illegible] | [illegible] | [illegible] | [illegible] | [illegible] | [illegible] | [illegible] |

| | | | | | | | | | |
|---|---|---|---|---|---|---|---|---|---|
| *TGFB1* | 19:41859051 (AC | 19: | 41859051 | A/AG | 0.01091 | INS | 5'UTR | | |
| *TGFB1* | 19:41859003 (TC | 19: | 41859003 | T/TG | 0.1255 | INS | 5'UTR | | |
| *TGFB1* | rs1349110564 | 19: | 41858963 | C/- | 0.03091 | DEL | 5'UTR | | |
| *TGFB1* | rs749452831 | 19: | 41858962 | G/GC | 0.005455 | INS | 5'UTR | | |
| *TGFB1* | rs759113717 | 19: | 41858937 | C/T | 0.001818 | SNP | Missense G->R | G5R | benign |
| *TGFB1* | rs1800470 | 19: | 41858921 | A/G | 0.4036 | SNP | Missense P->L | L10P | benign |
| *TGFB1* | rs1800471 | 19: | 41858876 | C/G | 0.06364 | SNP | Missense R->P | R25P | benign |
| *TGFB1* | 19:41858873 (C) | 19: | 41858873 | G/C | 0.01273 | SNP | Missense P->R | P26R | benign |
| *TGFB1* | rs199758510 | 19: | 41858864 | C/A | 0.001818 | SNP | Missense G->V | G29V | possibly damaging |
| *TGFB1* | 19:41858762 (CC | 19: | 41858762 | C/CG | 0.007273 | INS | Frameshift | | |
| *TGFB1* | rs200230522 | 19: | 41858739 | C/T | 0.001818 | SNP | Missense G->S | G71S | benign |
| *TGFB1* | 19:41858728 (CC | 19: | 41858728 | C/CG | 0.001818 | INS | Frameshift | | |
| *TGFB1* | 19:41850658 (-) | 19: | 41850658 | G/- | 0.01455 | DEL | Frameshift | | |
| *TGFB1* | 19:41848140 (-) | 19: | 41848140 | C/- | 0.01091 | DEL | Frameshift | | |
| *TGFB1* | rs55659002 | 19: | 41847943 | G/- | 0.007273 | DEL | intron | | |
| *TGFB1* | rs1800472 | 19: | 41847860 | G/A | 0.03818 | SNP | Missense T->I | T263I | benign |
| *TGFB1* | rs188080621 | 19: | 41847770 | C/G | 0.001818 | SNP | Intron | | |
| *TGFB1* | rs11466334 | 19: | 41847737 | G/A | 0.001818 | SNP | Intron | | |
| *TGFB1* | rs8179181 | 19: | 41838206 | G/A | 0.2527 | SNP | Intron | | |
| *TGFB1* | rs200550755 | 19: | 41838016 | C/T | 0.001818 | SNP | Intron | | |
| *TGFB1* | rs756877212 | 19: | 41836225 | T/C | 0.001818 | SNP | downstream | | |
| *TGFB1* | rs1030488680 | 19: | 41835164 | C/T | 0.001818 | SNP | downstream | | |
| *TGFB1* | rs117064611 | 19: | 41834818 | G/A | 0.02364 | SNP | downstream | | |
| *TGFB1* | rs143386142 | 19: | 41834559 | C/T | 0.001818 | SNP | downstream | | |
| *TGFB1* | rs1017131395 | 19: | 41834503 | C/G | 0.001818 | SNP | downstream | | |
| *TGFB1* | rs12983047 | 19: | 41834499 | A/G | 0.1564 | SNP | downstream | | |
| *TGFB1* | rs904198258 | 19: | 41834248 | C/T | 0.001818 | SNP | downstream | | |
| *TGFB1* | rs545612006 | 19: | 41834243 | G/A | 0.005455 | SNP | downstream | | |
| *TGFB1* | rs73045282 | 19: | 41833233 | G/A | 0.05636 | SNP | downstream | | |
| *TGFB1* | rs201781533 | 19: | 41833192 | G/GCA | 0.003636 | INS | downstream | | |
| *TGFB1* | rs10417924 | 19: | 41833167 | C/T | 0.2255 | SNP | downstream | | |
| *TGFB1* | rs142023663 | 19: | 41833151 | G/A | 0.001818 | SNP | downstream | | |
